# Subphenotyping of COVID-19 patients at pre-admission towards anticipated severity stratification: an analysis of 778 692 Mexican patients through an age-gender unbiased meta-clustering technique

**DOI:** 10.1101/2021.02.21.21252132

**Authors:** Lexin Zhou, Nekane Romero, Juan Martínez-Miranda, J Alberto Conejero, Juan M García-Gómez, Carlos Sáez

## Abstract

We apply a meta-clustering technique to discover age-gender unbiased COVID-19 patient subphenotypes based on phenotypical before admission, including pre-existing comorbidities, habits and demographic features, to study the potential early severity stratification capabilities of the discovered subgroups through characterizing their severity patterns including prognostic, ICU and morbimortality outcomes. We used the Mexican Government COVID-19 open data including 778,692 SARS-CoV-2 population-based patient-level data as of September 2020. The meta-clustering technique consists of a two-stage clustering approach combining dimensionality reduction and hierarchical clustering: 56 clusters from independent age-gender clustering analyses supported 11 clinically distinguishable meta-clusters (MCs). MCs 1-3 showed high recovery rates (90.27-95.22%), including healthy patients of all ages; children with comorbidities alongside priority in medical resources; and young obese smokers. MCs 4-5 showed moderate recovery rates (81.3-82.81%): patients with hypertension or diabetes of all ages; and obese patients with pneumonia, hypertension and diabetes. MCs 6-11 showed low recovery rates (53.96-66.94%): immunosuppressed patients with high comorbidity rate; CKD patients with poor survival length and recovery; elderly smokers with COPD; severe diabetic elderly with hypertension; and oldest obese smokers with COPD and mild cardiovascular disease. Group outcomes conformed to the recent literature on dedicated age-gender groups. These results can potentially help in the clinical patient understanding and their stratification towards automated early triage, prior to further tests and laboratory results are available, or help decide priority in vaccination or resource allocation among vulnerable subgroups or locations where additional tests are not available.

Code available at: https://github.com/bdslab-upv/covid19-metaclustering

## Introduction

In Mexico, mid-January 2020 reported the first cases of COVID-19. In early March 2020, the novel severe acute respiratory syndrome coronavirus 2 (SARS-CoV-2) was declared by the World Health Organization as a pandemic^1^. As of August 13th, 2020, a total of 20,439,814 confirmed cases of coronavirus disease 2019 (COVID-19) have been reported to the World Health Organization, and 744,385 lives have been lost^2^.

The COVID-19 pandemic has led to an unprecedented global healthcare challenge for both medical institutions and researchers. They have been making a huge effort to describe specific COVID-19 risk factors association, severity outcomes, and also personalized therapeutic options for COVID-19 patients being yet under investigation^3,4,5^. Recognizing different COVID-19 subphenotypes and their severity characterization may assist clinicians during the clinical course and research efforts. However, the availability of information to investigate such subphenotypes and consequent decision making often varies both according to the stage at which patients are in the COVID-19 clinical workflow –e.g., before admission, at admission, or during hospitalization– and according to the hospital access possibilities –e.g., hospitalized versus ambulatory patients–, especially in locations where hospitalization is difficult. In addition, the patient age and gender entail a potential correlation between subgroup characterization and their severity characterization which requires prudent usage in ML models.

Several studies have suggested potential COVID-19 subphenotypes, mainly within specific comorbidities such as pulmonary diseases or diabetes^6,7^ or related to distinct genetic variants^8^. However, the Mexican population has its particularity due to a high prevalence of comorbidities, like hypertension, diabetes –a leading cause of death in 2020^9^– and obesity, which is leading the population to a undesirable risks for severe coronavirus outcomes than many other high-income countries^10^. Since distinct target population often present heterogeneous clinical characterization and severity outcomes, it remains crucial a transparent understanding regarding the characterization of COVID-19 subphenotypes in Mexican patients to help anticipate individuals’ prognostic outcomes if one gets infected and evaluate subphenotypic severity presentations.

We describe the results of an unsupervised Machine Learning (ML) meta-clustering approach to identify potential subphenotypes of COVID-19 patients in Mexico based on previously existing comorbidities, habits and demographic features (i.e., age and gender). Stratification on gender and age groups was included for three primary reasons: (1) to reduce potential ML model’s biases in representing the best the majority (e.g. the young adults) but not underrepresented groups (e.g. children and elderly)^11^; (2) to reduce potential confounding factors from age and gender which are highly correlated with comorbidities, habits and mortality -i.e. age-gender clusters may help reveal more well-detailed patterns and phenotypical description-; and (3) to reduce interpretation biases, e.g. if one healthy cluster presents a mortality rate of 98.5% but includes patients from all ages, this specific mortality rate may vary across two patients from the same cluster whose age differ significantly (e.g. children versus adults). See section 1 of supplementary material for further details.

By using a population-based cohort of more than 700,000 patient-level cases, this is probably the largest cluster analysis about coronavirus patient-level cases to date. Other studies proposed unsupervised ML methods for aggregated population data^12^, CT image analyses^13,14^, molecular-level clustering^15^, or coronavirus-related scientific texts^16^. Several studies provided to date results from unsupervised ML on patient-level epidemiological data^17,18,19,20,21,22,23,24^. To our knowledge, however, none characterized age-gender subphenotypes, nor aimed to a population-based study with solely the phenotypical information available at pre-admission towards automated risk stratification, and neither characterized the Mexican population that is generally more vulnerable due to its particularity in a high prevalence of comorbidities.

Performing an accurate triage upon admission, but especially in ambulatory settings, is often challenging, significantly depending on the patient information available to the physicians. This work, therefore, aims to characterize age-gender COVID-19 subphenotypes that may potentially establish target groups for triage systems to assist clinicians in efficiently allocating limited resources and prioritize vaccination among subgroups that are more vulnerable when they get infected during the pandemic. As these subphenotypes are based on easily available data, such as previous disease and lifestyle habits rather than COVID-19 related symptoms (e.g., fever and nausea), vital signs and biomarkers that are not often available in the first days of COVID-19 infection or difficult to obtain due to limited resources, our work therefore could support early triage prior to further tests and laboratory results and even provide guidance in areas where such tests are not available.

## Results

### Age-gender cluster analysis

After evaluating the stratified clustering results, we selected the following number of clusters (k) for each specific age-gender group: <18-Male: k=5, <18-Female: k=4, 18-49-Male: k=7, 18-49-Female: k=7, 50-64-Male: k=9, 50-64-Female: k=8, >64-Male: k=8, >64-Female: k=8. This resulted in 56 age-gender clusters in total. Supplementary Table 5 describes the number of individuals for each age-gender cluster. By taking PCA scores, fed by the comorbidities and habits ratios of each group, we performed the second-stage meta-clustering analysis and established 11 clinically distinguishable MCs.

Figure 1 describes the relationships among different comorbidities and habits of the original 56 age-gender clusters through the first two principal components (Figure 1A), also provides the correspondence to their assigned MCs (Figure 1B) and their LOESS delineations for distinct severity outcomes (Figure 1C to 1H).

**Figure 1.**
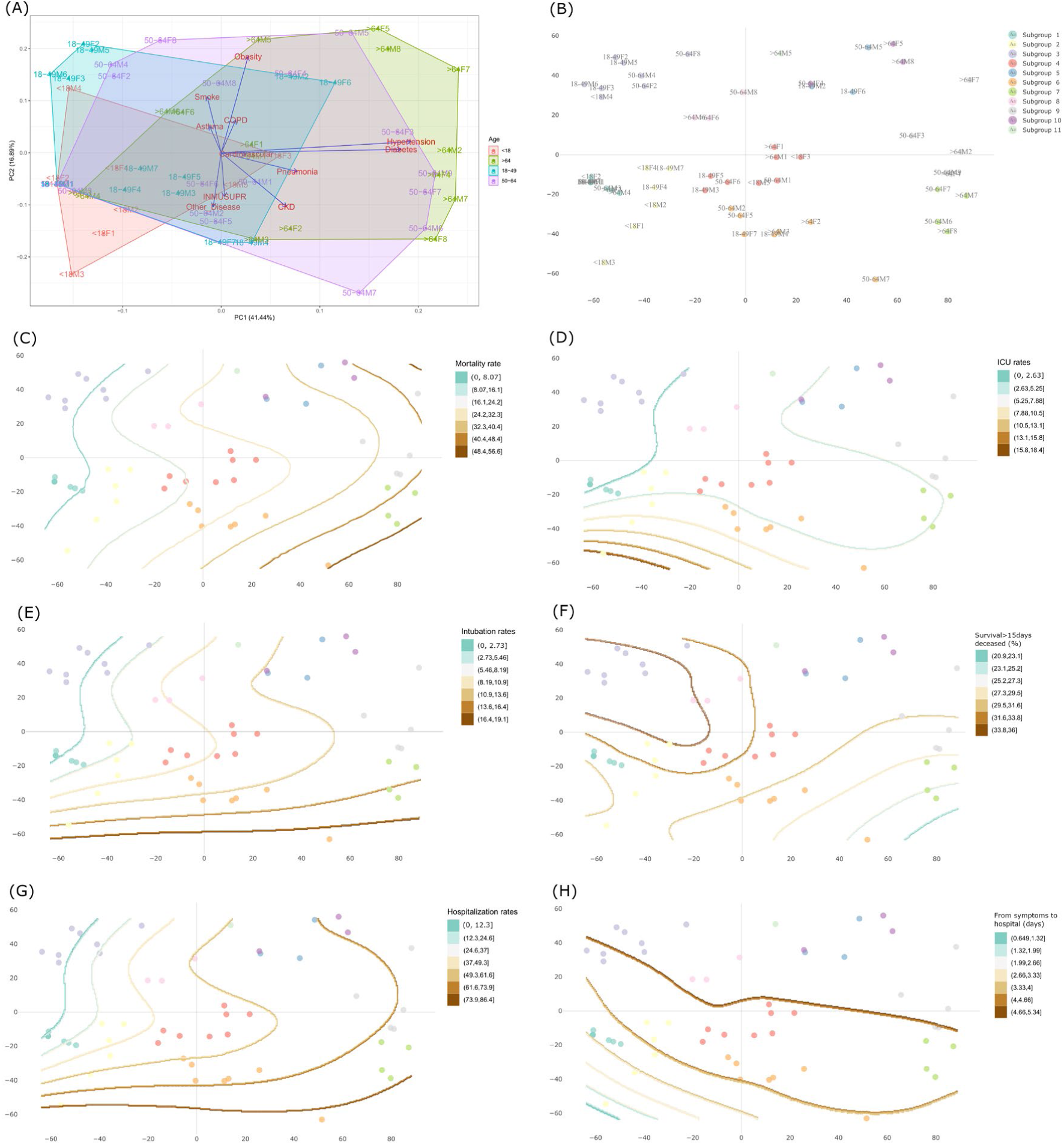
Principal component analysis (PCA) of the 56 age-gender clusters, meta-clustering results and LOESS-based severity delineations. (A) PCA from 56 age-gender stratified clusters; (B) scatterplot of the eleven MCs defined from the 56 clusters. (C-H) LOESS scatterplots regarding the severity outcomes of the eleven MCs among the 56 computed clusters. The LOESS models delineate seven severity ranges for each outcome including (C) mortality, (D) ICU admission, (E) intubation, (F) survival at 15 days among deceased patients, (G) hospitalization, and (H) days from symptoms to hospitalization. All the scatter plots share coordinates. Each subgroup is denoted using the following abbreviation: [AgeGroup][Gender][ClusterID]. Mexico, January 13–September 30, 2020.

The 56 clusters PCA analysis uncovered noticeable patterns and characterization among clusters of different ages in both genders. Young adults are prone to asthma and smoking habit; whereas the elderly was prone to many comorbidities such as hypertension, diabetes, obesity, COPD, pneumonia, and CKD. The results also show that obesity and smoking habit –both positively correlated– are strongly separated from immunosuppression and other diseases –both positively correlated–, implying these two pairs of features are negatively correlated in the studied data subgroups.

The LOESS models show that children took fewer days from presenting symptoms to hospitalization, showing higher ICU, intubation, and hospitalization rates than adults with similar conditions (Figure 1D, E, G, H). In contrast, MC3 –young obese cluster with moderate asthma and smoke rates– behaved inversely, implying that children, under similar clinical conditions, may receive priority regarding medical attention.

Inspecting the relationship between the PCA and LOESS models shows that CKD is significantly associated with a shorter survival length among deceased patients and an increase in intubation rates (Figure 1E, D). Mortality constantly increases from children to the elderly, but the most severe zones are inclined toward pneumonia, CKD, and COPD (Figure 1C) independently of the age groups.

Figure 2 describes and quantifies the features of the 56 age-gender clusters and relates them to their MC, and highlights relevant patterns through simultaneously ordering rows and columns through a biclustering technique^25^. Figure 2 reinforces that the children have a faster time from symptoms to hospitalization and are prone to ICU admission despite presenting similar clinical condition than adults (e.g., cluster <18M3 versus 50-64F5). Regarding gender discrepancy, females showed a better Recovery Rate (RR) despite presenting similar clinical conditions than males (e.g., >64M1 versus >64F1).

**Figure 2.**
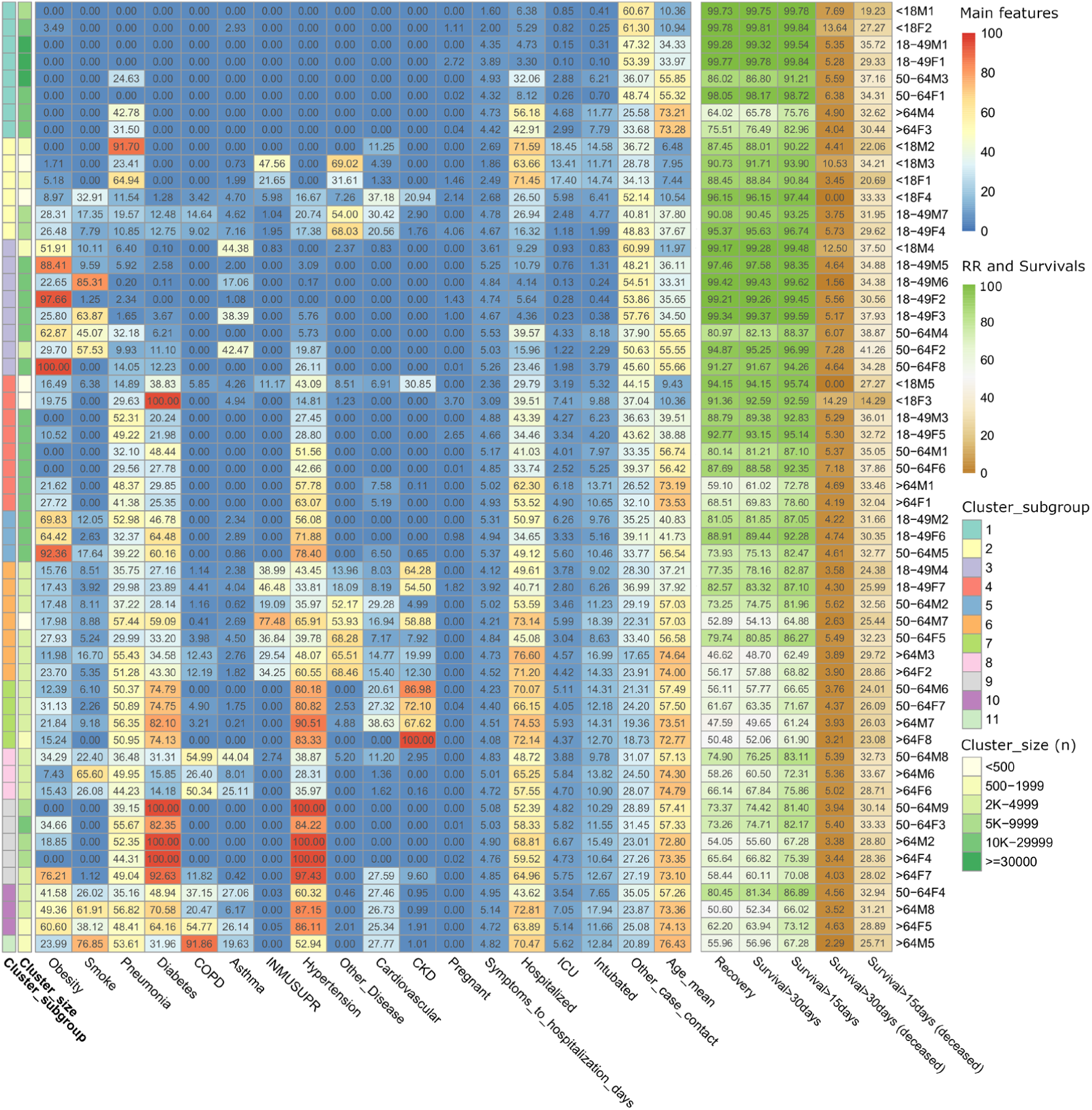
Heatmap showing the quantified characteristics among 56 of each age-gender specific cluster of the eleven MCs, the size of each cluster (n) was categorized into 6 ranges. Mexico, January 13–September 30, 2020.

The phenotypes, demographic features and outcomes of each cluster group can be fully explored at http://covid19sdetool.upv.es/?tab=mexicoGov.

### Epidemiological description of the 11 meta-clusters

Table 1 represents the quantified features of the 11 resultant MCs. Table 2 summarizes the 11 resultant MCs’ main features. Next, we describe the clinically distinguishable main epidemiological findings for each MC.

**Table 1.** The distribution of age, features and comorbidity with the quantitative description of demographic features, treatment, and epidemiological characteristics among eleven MCs. In these results, we applied arithmetic mean presuming that each age-gender cluster is representative to its population. Thus, the size (n) of each age-gender cluster was ignored. Mexico, January 13–September 30, 2020.

|  | MC1 | MC2 | MC3 | MC4 | MC5 | MC6 | MC7 | MC8 | MC9 | MC10 | MC11 |
| --- | --- | --- | --- | --- | --- | --- | --- | --- | --- | --- | --- |
| No. of age-gender clusters (n total = 56) | 8 | 6 | 8 | 8 | 3 | 7 | 4 | 3 | 5 | 3 | 1 |
| **Demographics (%)** |  |  |  |  |  |  |  |  |  |  |  |
| Age ($\bar{x}$) | 43.4 | 18 | 39.8 | 44.8 | 46.4 | 56.3 | 65.3 | 68.7 | 66.8 | 68.2 | 76.4 |
| Female | 50 | 50 | 50 | 50 | 33.33 | 42.86 | 50 | 33.33 | 60 | 66.67 | 0 |
| **Age range (%)** |  |  |  |  |  |  |  |  |  |  |  |
| <18 | 25 | 66.67 | 12.5 | 25 | 0 | 0 | 0 | 0 | 0 | 0 | 0 |
| 18-49 | 25 | 33.33 | 50 | 25 | 66.67 | 28.57 | 0 | 0 | 0 | 0 | 0 |
| 50-64 | 25 | 0 | 37.5 | 25 | 33.33 | 42.86 | 50 | 33.33 | 40 | 33.33 | 0 |
| >64 | 25 | 0 | 0 | 25 | 0 | 28.57 | 50 | 66.67 | 60 | 66.67 | 100 |
| Pregnancy (%) | 0.49 | 1.28 | 0.3 | 0.8 | 0.33 | 0.26 | 0.01 | 0 | 0.01 | 0 | 0 |
| **Habits (%)** |  |  |  |  |  |  |  |  |  |  |  |
| Obesity | 0.44 | 11.78 | 59.88 | 12.01 | 75.54 | 18.89 | 20.15 | 19.05 | 25.94 | 50.51 | 23.99 |
| Smoke | 0 | 9.67 | 34.09 | 0.8 | 10.77 | 8.1 | 4.38 | 38.03 | 0.22 | 42.02 | 76.85 |
| **Comorbidities (%)** |  |  |  |  |  |  |  |  |  |  |  |
| Diabetes | 0 | 4.42 | 4.5 | 39.06 | 57.14 | 35.62 | 76.44 | 20.45 | 95 | 61.23 | 31.96 |
| COPD | 0 | 4.51 | 0 | 0.73 | 0 | 5.1 | 2.03 | 43.91 | 2.36 | 37.46 | 91.86 |
| Asthma | 0.37 | 3.2 | 18.17 | 1.15 | 2.03 | 2.69 | 0.49 | 25.72 | 0.08 | 19.79 | 19.63 |
| INMUSUPR | 0 | 13.03 | 0.1 | 1.4 | 0 | 40.38 | 0 | 0.91 | 0 | 0.03 | 0 |
| Hypertension | 0 | 9.13 | 7.59 | 41.15 | 68.79 | 46.79 | 83.71 | 34.38 | 96.33 | 77.86 | 52.94 |
| Other Disease | 0 | 38.32 | 0.3 | 1.22 | 0 | 48.63 | 1.85 | 1.73 | 0 | 0.82 | 0 |
| Cardiovascular | 0 | 17.52 | 0.1 | 2.46 | 2.17 | 14.25 | 21.64 | 4.73 | 5.52 | 26.51 | 27.77 |
| CKD | 0 | 4.27 | 0 | 3.87 | 0.22 | 31.84 | 81.67 | 1.04 | 1.92 | 1.28 | 1.01 |
| **Treatment (%)** |  |  |  |  |  |  |  |  |  |  |  |
| Hospitalized | 19.87 | 46.08 | 14.15 | 42.22 | 44.91 | 58.56 | 70.72 | 57.17 | 60.8 | 60.11 | 70.47 |
| ICU | 1.59 | 9.82 | 1.23 | 4.48 | 5.06 | 4.01 | 4.87 | 4.81 | 5.56 | 5.24 | 5.62 |
| INTUBATED | 3.44 | 9.03 | 2.18 | 7.9 | 8.46 | 12.12 | 13.38 | 11.5 | 12.13 | 12.42 | 12.84 |
| Pneumonia (%) | 12.36 | 37 | 9.08 | 37.18 | 41.52 | 42.44 | 52.14 | 43.55 | 48.1 | 46.8 | 53.61 |
| Recovery (%) | 90.27 | 91.37 | 95.22 | 82.81 | 81.3 | 66.94 | 53.96 | 66.43 | 64.95 | 64.42 | 55.96 |
| Survival>15days (%) | 93.46 | 93.73 | 97.01 | 88.39 | 87.27 | 76.34 | 65.37 | 77.1 | 75.26 | 75.34 | 67.28 |
| Survival>30days (%) | 90.74 | 91.8 | 95.5 | 83.74 | 82.14 | 68.26 | 55.71 | 68.2 | 66.33 | 65.88 | 56.96 |
| Survival>15days, deceased (%) | 30.76 | 28.64 | 36.21 | 31.09 | 31.59 | 28.46 | 24.8 | 31.7 | 29.73 | 31.01 | 25.71 |
| Survival>30days, deceased (%) | 6.61 | 4.64 | 5.93 | 5.79 | 4.52 | 4.2 | 3.82 | 5.26 | 4.04 | 4.24 | 2.29 |
| From Symptoms to Hospital days ($\bar{x}$) | 3.78 | 3.2 | 4.87 | 4.37 | 5.21 | 4.48 | 4.3 | 4.85 | 4.92 | 4.94 | 4.82 |
| Other case contact (%) | 45.84 | 40.23 | 51.18 | 36.6 | 36.04 | 27.39 | 20.9 | 27.88 | 27.56 | 28 | 20.89 |

**Table 2.**
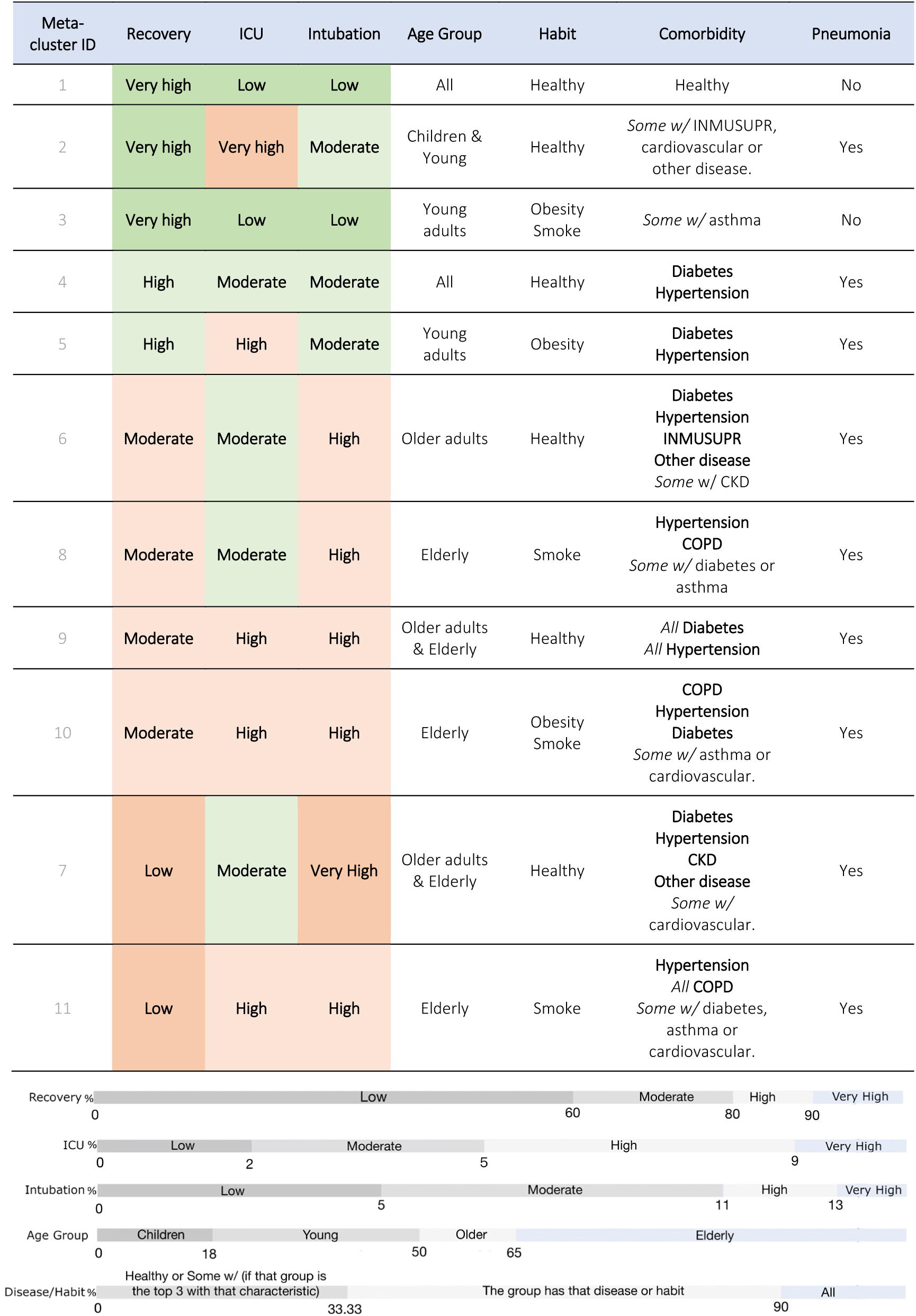
Main features of the 11 resultant meta-clusters, sorted by recovery. The thresholds for the different severity outcomes and input variable categories are displayed below. Mexico, January 13–September 30, 2020.

MC1 includes two healthiest clusters per each age-gender group, with a very high RR (90%). Most deceased patients in MC1 with pneumonia are older patients (Figure 2). MC2 includes children and young individuals (mean age 18) with healthy habits and little incidence of relevant diseases (13% immunosuppression, 17% cardiovascular disease, 4% CKD), albeit RR is very high (91%). Besides, MC2 also holds the highest ICU admission rate (9%), driven by three children clusters whose ICU rate vary from 13.41% to 18.45%. MC3 includes young adults (mean age 40) with significant obesity, smoking, a little incidence of other diseases and very high RR (95%). Despite the similarly high RRs in MC1 to 3, MC1 and MC3 show a low incidence of pneumonia, MC2 has 1/3 of pneumonia patients.

MC4 includes individuals of all ages with healthy habits but, unlike MC1, most patients in MC4 have hypertension (41%) or diabetes (39%), but not both simultaneously. MC5 includes young adults with obesity (75%), diabetes (57%) and/or hypertension (69%). Despite of this dissimilarity, MC4 and 5 still have similarly high RRs, of approximately 80%. From MC4 onwards, all MCs have from 40 to 50% incidence of pneumonia as of case reporting; what does not exclude the possibility that some patients developed pneumonia days after. Noteworthy, in groups 4 to 11 more than 70% of deceased patients were diagnosed with pneumonia.

The RRs from MC6 and 8-10 are similar (64-67%). MC6 includes older adults with no obesity nor smoking, but with frequent diseases including diabetes, hypertension, immunosuppression or other. MC8 includes elderly with smoking habit, plus hypertension (34%) and/or COPD (44%), two smoking-related diseases. Similarly, MC10 includes elderly with obesity (50%) or smoking habit (42%), who also suffer from COPD (37%), but with a much higher incidence of diabetes (61%) and hypertension (78%). MC9 contains older adults and elderly with both diabetes (95%) and hypertension (96%).

MC7 and 11 hold the lowest RRs (54% and 56%). MC7 includes older adults and elderly with common diseases –diabetes, hypertension and cardiovascular disease– plus CKD (81%). CKD stands out as the differential factor with similar MCs with low RRs, such as 6 or 9. MC11 is similar to 8 and 10; the key differences are the higher prevalence of smoking (78%, which doubles the former) and COPD (almost all patients, 91%), and a mean age eight years older (76 vs. 68 years). In addition, MCs that include older obese patients with smoking habits –MC8, 10, and 11– have significantly higher COPD and cardiovascular incidence, an association that does not occur with the young smokers –MC3.

### Variability among states and types of clinical institution

Regarding state variability, half of the Mexican states are prone to a higher probability in healthy clusters with better RR, lower hospitalization, ICU, and Intubation rates among each age-gender group (Figure 3A, e.g., 18F2, 18M1, 18-49F1, 18-49M1, 50-64F1) and MCs (Figure 3C), whereas another half behave inversely. Hidalgo, Baja California and Morelos represent healthiest groups as a contrast with Oaxaca, Coahuila de Zaragoza and Durango that represent the less healthy. Surprisingly, Mexico City showed significantly higher probability in healthier clusters than State of Mexico, albeit the population of their main urban areas are close, and both have similar resources and economic development level.

**Figure 3.**
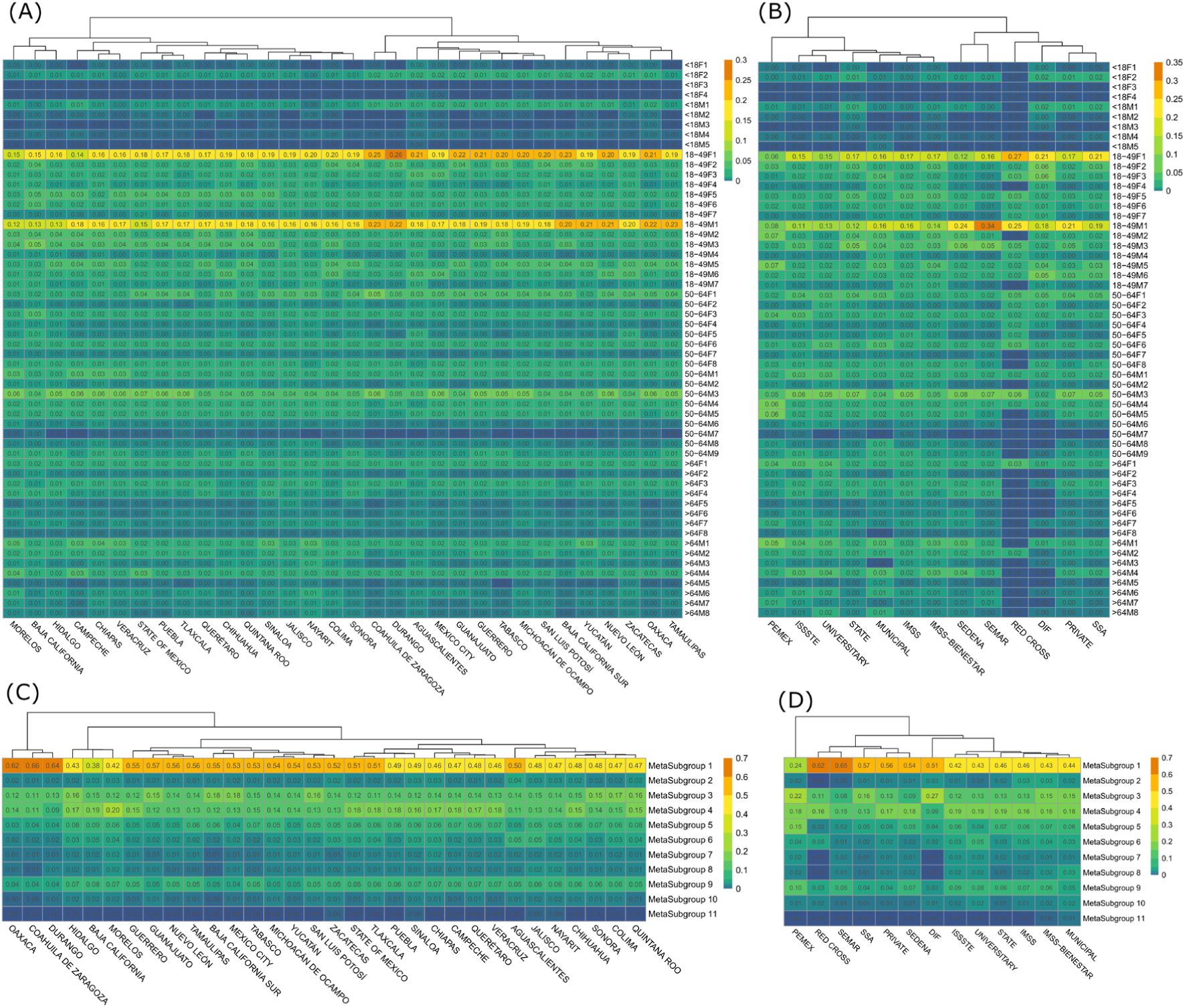
Heatmaps of the probability distribution of the 56 age-gender specific clusters (A, B) and eleven MCs (C, D) for each Mexican state where patients received the treatment or medical attention (A, C) and each type of clinical institution (TCI) (B, D). Rows represent the clusters and columns represent the states and TCI. Columns are arranged according to a hierarchical clustering on their values. Note that we compared the clusters’ distribution within each age range to circumvent any correlation or association with comorbidities and habits. Mexico, January 13–September 30, 2020. Abbreviations: DIF, National System for Integral Family Development; IMSS, Mexican Institute of Social Security; ISSSTE, Institute for Social Security and Services for State Workers; PEMEX, Mexican Petroleum Institution; SEDENA, Secretariat of the National Defense; SEMAR, Secretariat of the Navy; SSA, Secretariat of Health.

Regarding TCI variability (Figure 3B, D), SSA, DIF, Private and Red Cross are prone to healthier young patients. This pattern occurred inversely in other TCIs, especially the Mexican Petroleum Institution, whose severe cluster probabilities are generally higher. The clinical institutions of the armed forces (SEMAR, SEDENA) were mostly healthy, intuitively with a higher probability of male patients. Noteworthy, among the three primary TCIs in Mexico, the public health system (SSA) is prone to mild-comorbidity and have relatively higher probabilities in healthy clusters among each age-gender groups, mostly in MC1 (57%) and 3 (16%); whereas in the two main social security systems (IMSS, ISSSTE) the situation is just the opposite.

## Discussion

To date, several reports used cluster analysis to describe heterogeneity or characterization in COVID-19 patient-level epidemiological data^17,18,19,20,21,22,23,24^. To our knowledge, none characterized population-based data (778,692 patients) and neither analyzed the Mexican population to find potential subphenotypes through cluster analysis on age-gender controlled patient strata. Age-gender unbiased COVID-19 characterization is crucial for a robust subphenotype description as well as a deeper understanding of the inter-patient disease patterns based on exclusively pre-existing comorbidities and lifestyle habits. Such characterization could assist clinicians in anticipating individuals’ risk if one gets infected, the poor outcomes, and morbimortality as well as providing bases to improve early triage systems with limited resources.

Previous literature have reported isolated risk factors and association of several severe disease progression. However, such information has a potential limitation in clinical decision-making progress. In this work, no single clinical variable or lifestyle habit was enough to characterize the COVID-19 subphenotypes; a typical phenomenon when the data has many categorical variables. This reflects the reality of clinical practice: patients do not usually fall into subgroups of “all good” or “all bad” outcomes, neither the patient outcomes are conclusive by one single variable. However, when considering the variables together, our study uncovered 11 clinically distinguishable MCs among 56 plausible age-gender clusters; these MC-defined subphenotypes alongside age-gender stratification may represent different disease mechanisms and outcome.

Each of the 11 MCs shows clinical consistency: their group outcomes can be potentially predicted from the proposed input variables, according to the literature published up to date. From an outcome perspective, a dividing line can be clearly drawn between MCs 1-5 and 6-11. While the former’s have RRs always over 80%, the latter’s overall survival never exceeds 70%. Several factors can explain these findings, mainly the age, habits, comorbidities. Since all MCs contain 30-60% of women within their input age-gender clusters, the association between gender and mortality is hard to see based only on MCs. However, age-gender cluster analysis showed clearly better outcomes in females despite presenting similar conditions than males; a phenomenon that also occurs between patients that have similar conditions from different age groups. Therefore, considering both age-gender clusters and meta-clusters is essential for a better characterization that reveals more relevant detailed information in COVID-19 subphenotypes.

Hereinafter, we discuss our results in accordance with both MCs and age-gender clusters and relate them with supporting literature to discuss clusters’ clinical consistency through the associated risk factors, including age, habits and comorbidities, as well as on patient provenance and types of clinical institution.

### Age

Two groups with very high RRs are MC2 and MC3, which contain children and young adults. Age may play a protective role against the disease for two reasons. First, as proven by MC3 versus all single-aged groups (MCs 6-11), pneumonia incidence is lower in young healthy groups; hence, good RR could be attributable to mild SARS-CoV-2 disease. Second, as shown by good RRs in MC2 –children with severe disease, response to treatment is probably also better at younger ages.

Besides, children –MC2– showed to receive priority regarding medical attention than adults with similar clinical conditions in Mexico. After discussion with Mexican clinicians, one explanation seems to be that in early ages, the decompensation or deterioration caused by a pulmonary disease is faster than adults and with a higher risk of death. While in adults, there is often some time margin to evaluate the patient condition’s evolution before intubation or ICU admission, but not for children. Furthermore, if besides the presence of pneumonia, the groups are defined by conditions such as CKD and cardiovascular issues, a child who already has those issues could be perceived as much higher risk/more vulnerable than an elderly. These results are supported by recent literature, a study with a small cohort from Madrid^26^ found 10% of 41 children with SARS-CoV-2 required ICU admission. Another study^27^ showed that severe COVID-19 can also happen in small children and adolescents, where risk factors for ICU admission included age younger than one-month, male sex, presence of lower respiratory tract infection signs and presence of a pre-existing medical condition.

Regarding the association between older age and outcomes, MC6 to 11 are exclusively composed of older adults and elderly with poor outcomes; except MC6 that includes 28.57% of young adults. However, overall survival cannot be explained only by age but along with the presence of comorbidities and habits: while MC11 shows the highest mortality and mean age, MC7 shows a similar RR with a mean age approximately ten years younger, similarly to those groups with better RRs. Besides, as widely described in literature^28^, older chronological age is not necessarily linked to higher mortality but physiological age. MC1 and 4 support this fact since, despite containing the same number of groups of each age, they show similar RRs (MC1 90.27%, MC4 82.81%) to those RRs of groups composed only of young adults with little incidence of previous disease (MC2 91.37%, MC3 95.22%) and those groups made of young adults with some frequent diseases, such as diabetes and hypertension (MC5 81.30%).

These findings support that, while a young age predisposes to mild disease^28,29^, habits and comorbidities may play a key role in predicting mortality in older patients with SARS-CoV-2 infection rather than considering only the chronological age itself. Noteworthy, the clustering for the individual age-gender groups with age >64 years, revealed that centenarians –individuals of over 100 years of age– repeatedly fell in the age-gender clusters with better outcomes. This fact conforms with the well-studied good health and low frailty scores^30^ of this subpopulation. Therefore, age along with presence of habits and comorbidities are key factors to explain the dividing line between “high”, “moderate” as well as the “low” RRs.

### Habits

The role of obesity and smoking as risk factors for severe disease are complex, since they are both associated with the development of many conditions (e.g. COPD^31^ or cardiovascular^32^). In our study, the influence of obesity seems to be clear by comparing MC4 and 5: albeit both show diabetes, hypertension and moderate RRs (81-82%), they differentiate in that MC4 includes patients of all ages (25%) without obesity and MC5 contains mostly young adults (66.7%) who suffer from obesity. This seems to suggest that obese young adults may behave as “older”, implying higher mortality^28,33^. However, we found just the opposite in young individuals without pre-existing comorbidities: MC1 and 3 have similar RRs albeit MC3 contains a significant number (59.27%) of obese patients or smokers.

These findings suggest the role of habits cannot be considered alone, but always along with age, comorbidities and duration of unhealthy habits. Our results found that smoking is risk factors for severe COPD and cardiovascular, primarily in older patients –MC8, 10, 11. Therefore, it results reasonable that the longer the time as a smoker, the greater the incidence of severe disease. In young patients, however, the evidence of smoking’s negative influence is not so straightforward. Some reviews have presented current smoking as a protective factor versus former smoking, while it is clearly a risk factor versus never smoking^34^. Our results show that groups gathering young smokers have RRs which are not inferior to age-matched non-smoking groups, as proven by the RR of MC3 (95.22%, 34% smokers) versus MC2 (91.37%, 9.7% smokers).

Regarding obesity, its influence is not so clear in older groups since all have a high ratio in certain comorbidity. Still, in young obese patients without comorbidity (18-49M5 and 18-49F2), obesity seems unrelated to mortality. In conclusion, when evaluating habits, considering the patient’s age and the unhealthy duration may help establish more useful prognostics and correlations.

### Comorbidities

Diabetes and hypertension showed the highest prevalence among the recorded comorbidities. Their prevalence seems to explain the decrease in RRs from over 90% in MC1, 3 to 81% in MCs 4-5, all of which are young adult groups. In older MCs (6-11) it results harder to evaluate independently since both diseases are present in nearly every group, not specifically characterizing any cluster except MC9 that represents older patients with both diseases simultaneously (>95%) alongside a low RR (66.43%). These accord with current literature that reported both diabetes and hypertension are independent risk factors for severe disease^28,35,36^.

Immunosuppressed patients fall mostly on MC6 –older adults with diabetes, hypertension, immunosuppression and other diseases. Noteworthy, immunosuppressed patients were not in the clusters with the lowest RRs. This conforms with some reports that described immunosuppression has not been confirmed as a relevant factor for disease severity, except for cancer patients^37,38^. MC6 also holds few CKD patients, a factor which has been studied as a key factor for disease progression^39,40^ and it may be the cause for the immunosuppression in this group (Odds Ratio: 9.65 95%CI [9.05-10.28]) according to the prevalence of immunosuppression of CKD patients versus non-CKD patients.

MC7 is characterised by the high prevalence of CKD and other diseases. Here, RR decreases roughly 10% compared with other severe subgroups. We found CKD highly associated with mortality and a shorter survival length. This accords with a report that revealed CKD was the factor that best explains mortality^41^, implying CKD patient could be vulnerable.

MC8 is similar to 10 and 11 to some extent since they all have COPD, where MC 11 gathering the most COPD patients (>90%). Most COPD patients are elderly with comorbidities with poor outcomes which conforms to several reviews that reported COPD patients with an increased risk of severe pneumonia and poor outcomes when they develop COVID-19^42,43^.

Cardiovascular disease is homogeneously distributed among groups, particularly on MC7, 10 and 11. Nowadays, cardiovascular disease may be a double-edged factor, since it is a proven risk factor for COVID-19 severity, but some of the treatments used, such as ACE inhibitors, have also proved to protect against severe infections from SARS-CoV-2^44,45^.

### State and Types of Clinical Institution

Reliable subphenotype characterization that reflects the geographical and healthcare settings from which they are ascertained is crucial^46^. To date, variability between Mexican states and TCIs regarding severity are rarely reported^47,48,49^, nor assessed for variability independently from age and gender. As an example, one state (e.g., Morelos) may show higher severity if it includes more elderly and male patients, but when we compare age-gender groups the results show that no severity difference exists in terms of probability within age-gender groups of the same age range.

The inter-state and TCI variability we found may be influenced by many factors such as the number and type –urban/rural– of population, sociocultural context, healthcare policy, the quantity of medical institutions and availability of resources, virus transmission level. Some states are more industrialized and more economical resources (e.g., Mexico City, Jalisco, the State of Mexico) than others (e.g., Oaxaca, Chiapas, Guerrero). The differences found between Mexico City and State of Mexico regarding healthy clusters distribution are hard to explain due to their proximity and similarities in the type of population and availability of medical resources.

One possible explanation for the differences in severity between social security institutions (IMSS and ISSSTE) and local public hospitals (SSA) is that SSA are administrated by the local states, and the resources among states often differ. This phenomenon could influence these institutions’ quality and resources to attend their populations. Another supportive explanation is that when SSA receives severe patients and have insufficient medical resources, these patients can be transferred to the IMSS COVID-19 facilities. Consequently, this may saturate IMSS and deplete the limited resources due to an increasing number of patients, making the distribution of resources harder. These results conform with previous studies showing that the risk of death for an average patient attending IMSS and ISSSTE is twice the national average and 3 times higher relative to the private clinical institution^47^.

The complex correlation between severity and state/TCI implies a crucial socioeconomic and healthcare resource level inequality. Thus, both considering state and TCI combined with MCs and age-gender clusters may help lead to a better subphenotype characterization.

### Limitations

As a possible limitation, we excluded patients confirmed after September 30 to avoid possible analysis disturbance on survival outcomes, which impeded us using the most recent data whose epidemiological characteristics could have changed to some degree. Furthermore, the dataset did not include additional relevant information about the patients who were discharged, readmitted, vaccinated and neither the duration of comorbidities and unhealthy habits. Further studies with population-based data regarding subphenotypes characterization among discharged patients who received post-surveillance or were readmitted or vaccinated population are highly needed.

In summary, the analysis of COVID-19 subphenotypes from the proposed two-stage cluster analysis produced clinically coherent models with discriminative characterization and explainability over age and gender. The resultant eleven MCs provide bases for a deep understanding of the epidemiological and subphenotypic characterization of COVID-19 patients based on pre-existing comorbidities, habits, demographic characteristics, and on patient provenance and types of clinical institution, as well as revealing the correlations between the above characteristics to anticipate the possible clinical outcomes of each patient with a specific profile. These unbiased subphenotypes may help establish target groups for automated stratification or triage systems to support clinicians with the early triage prior to further tests and laboratory results, especially in those areas where such tests are not available, and prioritize vaccination among the general population as well as provide bases in planning personalized therapies or treatments. For example, a CKD patient could be classified into subgroups –MC6, 7– distinguished by pervasive differences in severity and comorbid patterns, and then compared with their inner age-gender groups whose profile coincide the most with our patient, enabling a personalized evaluation of the patient’s prognostic outcomes and severity presentation.

Combining age-gender stratification and meta-clustering technique successfully revealed well-detailed informative findings and are potential for designing a novel data-driven model for the stratification of COVID-19 patients with the information available at pre-admission, or even to anticipate outcomes on the general population. Besides, the results shed light on robust conclusions about association and causality between subphenotypic presentation and clinical outcomes. Future study can explore the treatment and vaccination implications toward providing guidance on clinical triage and customize therapy, and also develop clinically robust subphenotype classification methodologies combined with the proposed two-stage cluster analysis. As the concern paid to efficient triage and personalized treatment increases, we facilitate further replicability of the study and generalization to other countries data by making available our experiment codes.

## Methods

### Data collection and processing

We used the publicly available COVID-19 Open Data by the Mexican Government^50^. As of 2 November 2020, the dataset comprises a total of 2,414,882 cases including demographic, comorbidities, habits, and prognosis patient-level data, for both positive and non-positive cases.

Figure 4 describes the study inclusion and exclusion criteria, and the data quality assessment process outcomes in a CONSORT-like flowchart. The final sample included 778,692 positive cases.

**Figure 4.**
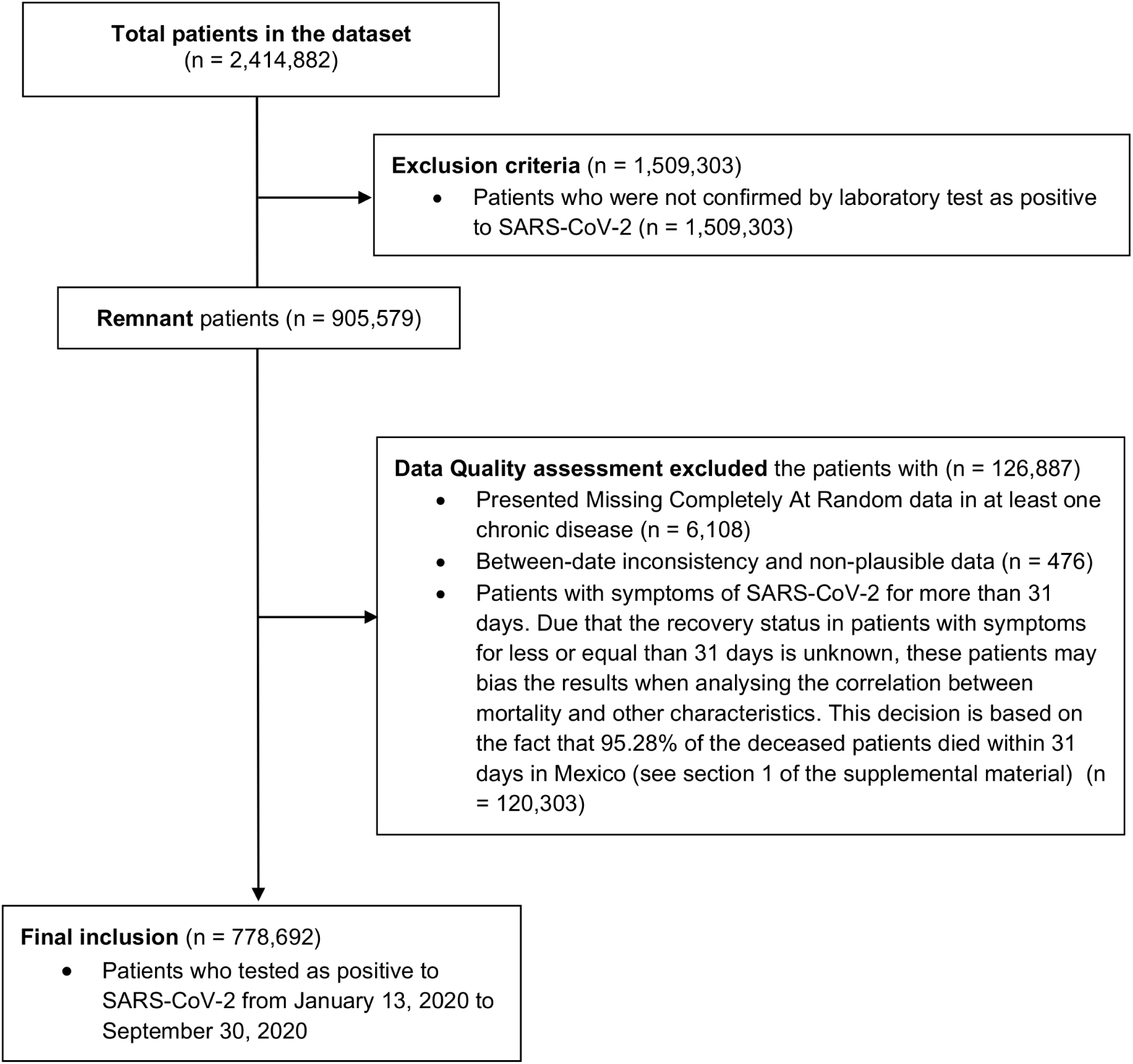
Dataset preprocessing flowchart.

We derived five outcome variables related with the prospective patient’s severity. First, the patient outcome, as deceased or not, from the date of death record. Second, the survival days since the date of symptoms. Third, the days from presenting symptoms to hospital admission. Lastly, we categorized the overall survival at 15 and 30 days after presenting symptoms.

After an assessment of potential temporal biases using temporal variability statistical methods^51^, and considering not significant temporal changes, we decided keeping the data from all the period of the study.

Table 3 shows the list of studied variables. The supplementary material, in sections 3 to 6, describes additional information on the data quality and variability analysis, the original dataset description, baseline characteristics of the COVID-19 patients alongside descriptive statistics in age-gender groups of the study sample, and pregnancy association in outcomes.

**Table 3.**
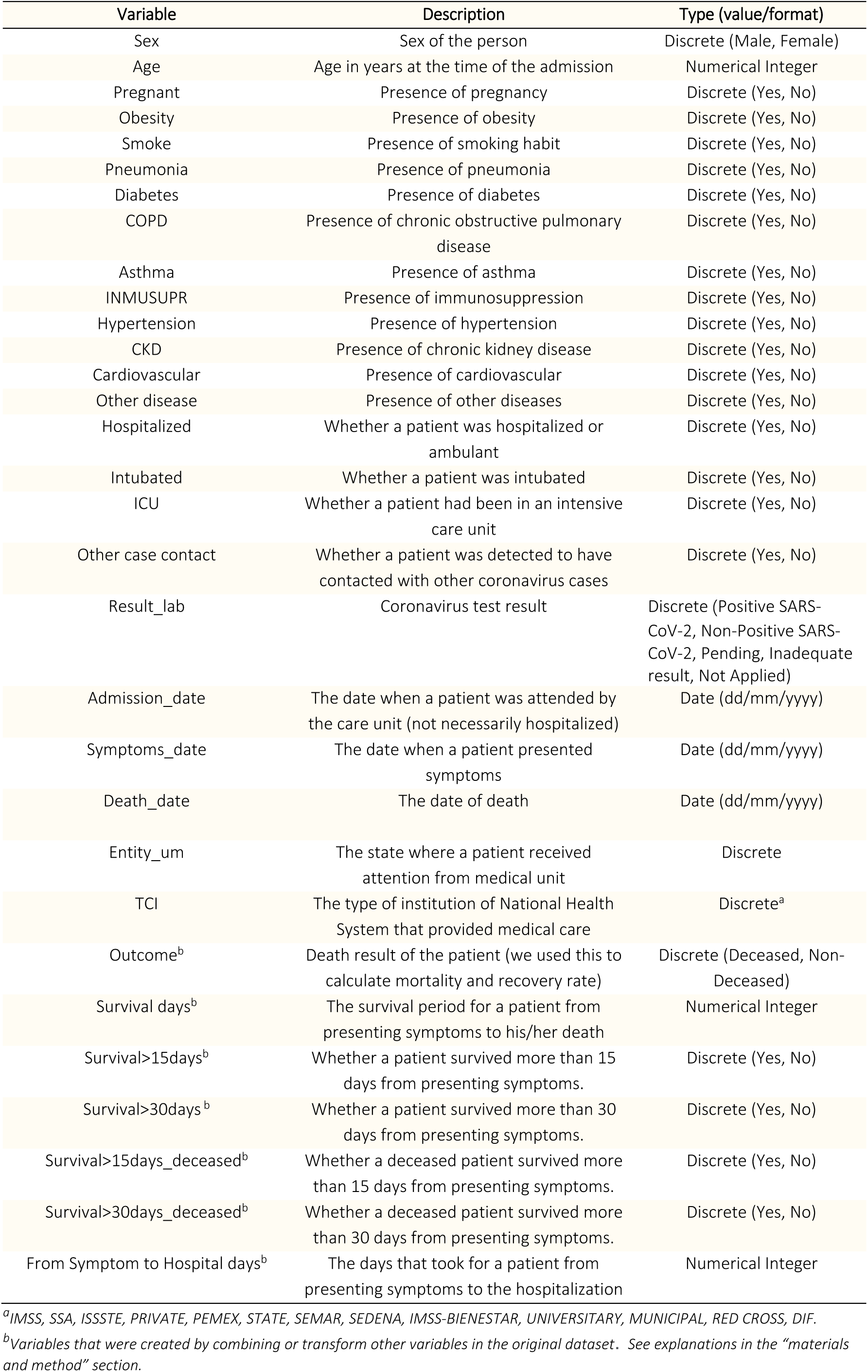
List of variables contained in the study case. Originally coded in Spanish, translated into English by the authors for this work. Abbreviations: COPD, chronic obstructive pulmonary disease; INMUSUPR, immunosuppression; CKD, chronic kidney disease; ICU, intensive care unit; TCI, type of clinical institution. Supplementary Table 2 describes the original dataset description.

| Variable | Description | Type (value/format) |
| --- | --- | --- |
| Sex | Sex of the person | Discrete (Male, Female) |
| Age | Age in years at the time of the admission | Numerical Integer |
| Pregnant | Presence of pregnancy | Discrete (Yes, No) |
| Obesity | Presence of obesity | Discrete (Yes, No) |
| Smoke | Presence of smoking habit | Discrete (Yes, No) |
| Pneumonia | Presence of pneumonia | Discrete (Yes, No) |
| Diabetes | Presence of diabetes | Discrete (Yes, No) |
| COPD | Presence of chronic obstructive pulmonary disease | Discrete (Yes, No) |
| Asthma | Presence of asthma | Discrete (Yes, No) |
| INMUSUPR | Presence of immunosuppression | Discrete (Yes, No) |
| Hypertension | Presence of hypertension | Discrete (Yes, No) |
| CKD | Presence of chronic kidney disease | Discrete (Yes, No) |
| Cardiovascular | Presence of cardiovascular | Discrete (Yes, No) |
| Other disease | Presence of other diseases | Discrete (Yes, No) |
| Hospitalized | Whether a patient was hospitalized or ambulant | Discrete (Yes, No) |
| Intubated | Whether a patient was intubated | Discrete (Yes, No) |
| ICU | Whether a patient had been in an intensive care unit | Discrete (Yes, No) |
| Other case contact | Whether a patient was detected to have contacted with other coronavirus cases | Discrete (Yes, No) |
| Result_lab | Coronavirus test result | Discrete (Positive SARS-CoV-2, Non-Positive SARS-CoV-2, Pending, Inadequate result, Not Applied) |
| Admission_date | The date when a patient was attended by the care unit (not necessarily hospitalized) | Date (dd/mm/yyyy) |
| Symptoms_date | The date when a patient presented symptoms | Date (dd/mm/yyyy) |
| Death_date | The date of death | Date (dd/mm/yyyy) |
| Entity_um | The state where a patient received attention from medical unit | Discrete |
| TCI | The type of institution of National Health System that provided medical care | Discrete<sup>a</sup> |
| Outcome<sup>b</sup> | Death result of the patient (we used this to calculate mortality and recovery rate) | Discrete (Deceased, Non-Deceased) |
| Survival days<sup>b</sup> | The survival period for a patient from presenting symptoms to his/her death | Numerical Integer |
| Survival>15days<sup>b</sup> | Whether a patient survived more than 15 days from presenting symptoms. | Discrete (Yes, No) |
| Survival>30days<sup>b</sup> | Whether a patient survived more than 30 days from presenting symptoms. | Discrete (Yes, No) |
| Survival>15days_deceased<sup>b</sup> | Whether a deceased patient survived more than 15 days from presenting symptoms. | Discrete (Yes, No) |
| Survival>30days_deceased<sup>b</sup> | Whether a deceased patient survived more than 30 days from presenting symptoms. | Discrete (Yes, No) |
| From Symptom to Hospital days<sup>b</sup> | The days that took for a patient from presenting symptoms to the hospitalization | Numerical Integer |

<sup>a</sup>IMSS, SSA, ISSSTE, PRIVATE, PEMEX, STATE, SEMAR, SEDENA, IMSS-BIENESTAR, UNIVERSITY, MUNICIPAL, RED CROSS, DIF.
<sup>b</sup>Variables that were created by combining or transform other variables in the original dataset. See explanations in the “materials and method” section.

### Meta-clustering methodology

We applied a two-stage subgroup discovery approach. In both stages, we used Ward’s minimum variance method with Euclidean squared distance^52^ to performed hierarchical clustering fed by a dimensionality reduction algorithm –Principal Component Analysis (PCA)^53^ or Multiple Correspondence Analysis (MCA)^54^– that took as input eleven variables including nine comorbidities –pneumonia, diabetes, COPD, asthma, INMUSUPR, hypertension, CKD, cardiovascular, and other diseases– alongside two unhealthy habits, namely obesity and smoking. In order to select the most representative PCA and MCA components to feed hierarchical clustering, we considered values with an eigenvalue higher than the average. Dimensionality reduction is known to help in the process of clustering by compressing information into a smaller number of variables, making unsupervised learning less prone to overfitting^55^, as well as to facilitate further visual analytics to prevent the potential ML black-box issue^56^.

In the first-stage, we applied individually hierarchical clustering analyses taking as input the MCA scores fed by comorbidities and habits at stratified groups according to gender and age (<18, 18-49, 50-64, and >64 years) to reduce potential biases and confounding factors, since age and gender are highly correlated with comorbidity, habits and mortality. Afterwards, we applied PCA and locally estimated scatterplot smoothing^57^ (LOESS) model on the resultant age-gender clusters’ features to visually explain their correlations and severity relationships. We created the cluster heatmap to help understand the characteristics of each age-gender cluster.

In the second-stage, in a wider perspective description of the population, we performed a hierarchical clustering again fed by PCA scores obtained via the resultant age-gender clusters taking as input their comorbidities and habits ratios. Then, we quantified the features of the resultant meta-clusters (MCs) representing via a table and also summarized these quantified features into a qualitative table to help interpret the main features of the resultant MCs.

For each subgroup analysis, we implemented cluster analyses from 2 through 12 clusters. The proper number of subgroups were obtained by combining a quantitative approach using Silhouette Coefficient^58^ –which measures the tightness and separation of the objects within clusters, reflecting how similar an object is to its own cluster compared to other clusters– and a qualitative cluster analysis audited by the authors of this work, including medical, health informatics and ML experts from Spain and Mexico. We first selected the group of clusters that showed relatively better Silhouette Coefficient values, then adjusted the number for the most reasonable and clinically distinguishable groups regarding clinical phenotypes. This process was supported by the pipelines and exploratory tool we developed in previous work^59^.

Finally, we performed source variability assessment^60^ using heatmaps to analyze the severity tendency among different data sources based on the clusters’ probability distribution between Mexican states and several types of clinical institution (TCIs) where patients received medical attention.

The data processing and analyses were performed using RStudio (version 3.6) and Python (version 3.8). Temporal and source variability –data quality analyses– were performed using the EHRtemporalVariability^51^ and EHRsourceVariability^60,61,62^ packages. Figure 5 summarizes the full methodology. The methods developed in this work are available in our GitHub repository https://github.com/bdslab-upv/covid19-metaclustering.

**Figure 5.**
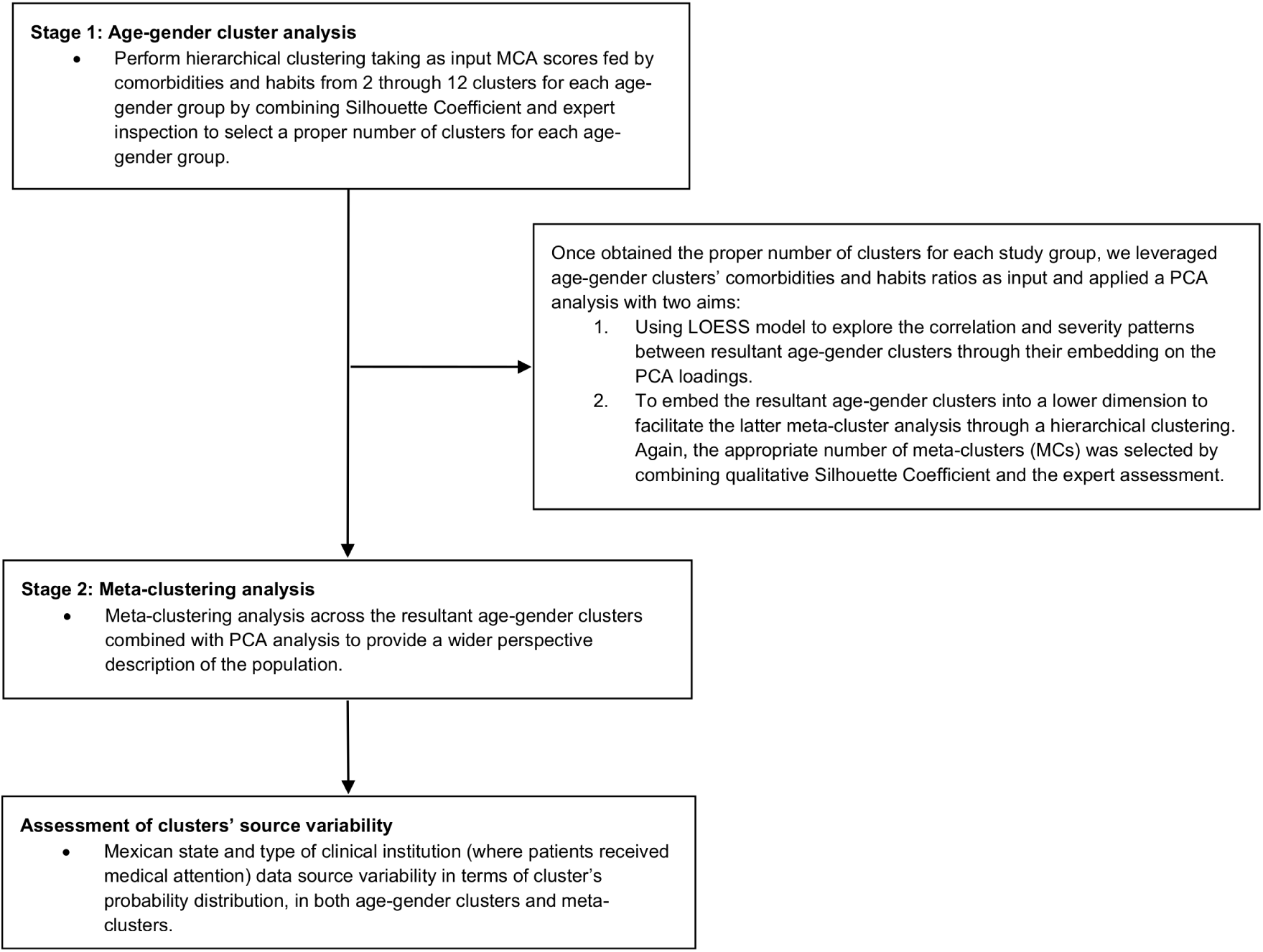
Research methodology flowchart.

## Data Availability

The studied sample is available in our GitHub repository.

https://github.com/bdslab-upv/covid19-metaclustering

## Abbreviations

COVID-19: coronavirus disease 2019
SARS-CoV-2: severe acute respiratory syndrome coronavirus 2
ML: Machine Learning
PCA: Principal Component Analysis
MCA: Multiple Correspondence Analysis
LOESS: locally estimated scatterplot smoothing
COPD: Chronic Obstructive Pulmonary Disease
CKD: Chronic Kidney Disease
INMUSUPR: Immunosuppression
ICU: Intensive Care Unit
RR: Recovery Rate
MC: Meta-Cluster
TIC: Types of Clinical Institution
DIF: National System for Integral Family Development
IMSS: Mexican Institute of Social Security
ISSSTE: Institute for Social Security and Services for State Workers
PEMEX: Mexican Petroleum Institution
SEDENA: Secretariat of the National Defense
SEMAR: Secretariat of the Navy
SSA: Secretariat of Health

## Data availability

The data of epidemiological and clinical patient-level open-source database in Mexico is publicly available at https://www.gob.mx/salud/documentos/datos-abiertos-152127 in Spanish. The English version and the studied sample of this dataset are available in our GitHub Repository https://github.com/bdslab-upv/covid19-metaclustering.

## Code availability

The codes that support the findings of this study are available at: https://github.com/bdslab-upv/covid19-metaclustering. The results from 2 through 12 clusters for both gender and age subgroups are available at http://covid19sdetool.upv.es/?tab=mexicoGov.

## Funding

This work was supported by Universitat Politècnica de València contract no. UPV-SUB.2-1302 and FONDO SUPERA COVID-19 by CRUE-Santander Bank grant: “Severity Subgroup Discovery and Classification on COVID-19 Real World Data through Machine Learning and Data Quality assessment (SUBCOVERWD-19)”.

## Acknowledgements

We sincerely thank the different types of clinical institutions and the Mexican government that have made a huge effort to make these data publicly available. We also thank the clinicians and epidemiologists from the Servicios de Salud de Nayarit for the useful discussions on specific aspects of the medical attention to hospitalized patients and the reporting of epidemiological data processes related to COVID-19. Furthermore, we would also like to thank Francisco Tomás García Ruiz for his valuable help in data visualization design.

## Authorship Statement

L.Z, C.S, J.M.G.G, J.A.C designed the research; L.Z, N.R, C.S, J.M.G.G, J.A.C, J.M.M conducted the research; L.Z, C.S processed and analyzed the data and performed the statistical analysis; all authors assessed the clinical consistency of the cluster analyses. L.Z, N.R, C.S drafted the manuscript; all authors: revised the manuscript critically; all authors approved the final manuscript.

## Competing interests

The authors declare no competing interests.

## Notes

### Competing Interest Statement

The authors have declared no competing interest.

### Author Declarations

Using Open Data from the Government of Mexico, terms available at: https://datos.gob.mx/libreusomx

### Summary of Updates

(1) Revise the article. (3) Supplemental files updated.

