## Appendix A-Supplemental Material for "Subphenotyping of COVID-19 patients at pre-admission towards anticipated severity stratification: an analysis of 778 692 Mexican patients through an age-gender unbiased meta-clustering technique"

[a]Biomedical Data Science Lab, Instituto Universitario de Tecnologías de la Información y Comunicaciones (ITACA), Universitat Politècnica de València (UPV), Camino de Vera s/n, Valencia 46022, España. [b]Instituto Universitario de Matemática Pura y Aplicada (IUMPA), Universitat Politècnica de València, Valencia, Spain. [c]CONACyT - Centro de Investigación Científica y de Educación Superior de Ensenada - CICESE-UT3, Mexico

†Senior authors

*Corresponding author

### Supplementary Material

### 1. The importance of cluster analyses with stratification on age-gender groups

This section describes why it is important to use cluster analyses with stratification on age-gender groups. Table 1 describes the quantified results of the 11 clusters found through hierarchical clustering using Ward's minimum variance method with Euclidean squared distance taking as input the PCA scores obtained via comorbidities and the habits.

For example, through observing cluster 1 –healthy without any previous condition and unhealthy habits– we may interpret that if the patient is a healthy person (no comorbidities nor unhealthy habits), his/her expected recovery rate will be 98.50 (95%CI; 98.46-98.54). However, this result may lead to interpretive biases since this recovery rate may vary between different age groups or even between male and female. This reflects that fact that the patient's outcome will vary depending on some underlying features, such as physiological ones (e.g., exercising routine, body strength, daily diet, immune response, vaccination record) that are generally hard to measure or unregistered.

This underlying bias was also verified in our main manuscript, where we found that despite having similar pre-existing conditions between children and adults, their overall clinical outcomes differ. The reason may be attributed to that in early ages, the decompensation or deterioration caused by a pulmonary disease is faster than adults and with a higher risk of death. While in adults, there is often some time margin to evaluate the patient condition's evolution before intubation or ICU admission, but not for children. Furthermore, if besides the presence of pneumonia, the groups are defined by conditions such as CKD and cardiovascular issues, a child who already has those issues could be perceived as much higher risk/more vulnerable than an elderly. We also found this bias between males and females with similar pre-existing conditions that we discussed in our main manuscript.

Therefore, we conclude that the analysis without including age-gender stratification but only a general cluster analysis for the entire population may produce interpretive biases and neither provide as much as well-detailed information and patterns that age-gender clusters can reveal.

**Table 1**. The distribution of age, features and comorbidity with the quantitative description of demographic features, treatment, and epidemiological characteristics among eleven clusters without age-gender stratification. Mexico, January 13–September 30, 2020.

| | Subgroup 1 | Subgroup 2 | Subgroup 3 | Subgroup 4 | Subgroup 5 | Subgroup 6 | Subgroup 7 | Subgroup 8 | Subgroup 9 | Subgroup 10 | Subgroup 11 |
|---|---|---|---|---|---|---|---|---|---|---|---|
| No. of individuals ($n_{total}$ = 778692) | 385318 | 76234 | 85338 | 61207 | 10265 | 3099 | 106269 | 18372 | 14769 | 2842 | 14979 |
| **Features (%\|x, CI 95%)** | | | | | | | | | | | |
| OBESITY | 0.00 (0.00-0.00) | 28.48 (28.16-28.81) | 100.00 (100.00-100.00) | 25.55 (25.21-25.90) | 30.45 (29.56-31.34) | 45.27 (43.52-47.03) | 0.00 (0.00-0.00) | 13.07 (12.59-13.56) | 6.57 (6.17-6.97) | 24.17 (22.60-25.75) | 51.04 (50.24-51.85) |
| SMOKE | 0.00 (0.00-0.00) | 0.00 (0.00-0.00) | 0.00 (0.00-0.00) | 75.16 (74.82-75.50) | 18.15 (17.40-18.89) | 24.07 (22.57-25.58) | 0.00 (0.00-0.00) | 2.84 (2.60-3.08) | 2.04 (1.82-2.27) | 12.03 (10.84-13.23) | 47.95 (47.15-48.75) |
| **Comorbidities (%\|x, CI 95%)** | | | | | | | | | | | |
| PNEUMONIA | 0.00 (0.00-0.00) | 64.36 (64.03-64.70) | 13.07 (12.85-13.30) | 10.01 (9.77-10.25) | 16.75 (16.02-17.47) | 60.31 (58.59-62.03) | 44.93 (44.63-45.23) | 49.07 (48.35-49.80) | 23.56 (22.87-24.24) | 43.60 (41.77-45.42) | 62.11 (61.33-62.88) |
| DIABETES | 0.00 (0.00-0.00) | 77.32 (77.02-77.62) | 7.31 (7.13-7.48) | 5.52 (5.34-5.70) | 8.61 (8.07-9.15) | 66.67 (65.01-68.33) | 22.22 (21.97-22.47) | 54.95 (54.23-55.67) | 15.27 (14.69-15.85) | 38.85 (37.05-40.64) | 68.71 (67.97-69.45) |
| COPD | 0.00 (0.00-0.00) | 0.00 (0.00-0.00) | 0.00 (0.00-0.00) | 2.43 (2.30-2.55) | 32.14 (31.23-33.04) | 56.02 (54.27-57.77) | 0.00 (0.00-0.00) | 0.55 (0.44-0.66) | 0.00 (0.00-0.00) | 14.71 (13.41-16.01) | 27.24 (26.53-27.95) |
| ASTHMA | 0.00 (0.00-0.00) | 0.00 (0.00-0.00) | 0.00 (0.00-0.00) | 26.42 (26.07-26.77) | 7.57 (7.06-8.08) | 8.39 (7.41-9.37) | 0.00 (0.00-0.00) | 0.45 (0.35-0.55) | 0.89 (0.74-1.05) | 7.14 (6.20-8.09) | 16.22 (15.63-16.81) |
| INMUSUPR | 0.00 (0.00-0.00) | 0.00 (0.00-0.00) | 0.00 (0.00-0.00) | 0.00 (0.00-0.00) | 1.95 (1.68-2.22) | 8.20 (7.23-9.16) | 0.00 (0.00-0.00) | 0.00 (0.00-0.00) | 34.68 (33.91-35.45) | 96.24 (95.54-96.93) | 0.00 (0.00-0.00) |
| HYPERTENSION | 0.00 (0.00-0.00) | 81.39 (81.12-81.67) | 15.73 (15.49-15.98) | 8.62 (8.39-8.84) | 17.09 (16.36-17.82) | 83.35 (82.04-84.66) | 32.85 (32.57-33.14) | 71.58 (70.93-72.23) | 18.69 (18.07-19.32) | 49.19 (47.35-51.03) | 81.01 (80.39-81.64) |
| OTHER DISEASE | 0.00 (0.00-0.00) | 0.00 (0.00-0.00) | 0.00 (0.00-0.00) | 0.00 (0.00-0.00) | 37.38 (36.44-38.32) | 24.81 (23.29-26.34) | 0.00 (0.00-0.00) | 14.83 (14.31-15.34) | 65.32 (64.55-66.09) | 54.36 (52.53-56.19) | 0.02 (0.00-0.04) |
| CARDIOVASCULAR | 0.00 (0.00-0.00) | 0.00 (0.00-0.00) | 0.00 (0.00-0.00) | 0.16 (0.13-0.20) | 31.05 (30.15-31.94) | 70.38 (68.77-71.99) | 0.00 (0.00-0.00) | 28.12 (27.47-28.77) | 3.23 (2.94-3.51) | 27.41 (25.77-29.05) | 21.24 (20.58-21.89) |
| CHRONIC KIDNEY DISEASE | 0.00 (0.00-0.00) | 0.00 (0.00-0.00) | 0.00 (0.00-0.00) | 0.00 (0.00-0.00) | 2.17 (1.89-2.45) | 48.63 (46.87-50.39) | 0.00 (0.00-0.00) | 58.41 (57.70-59.13) | 3.55 (3.26-3.85) | 45.64 (43.81-47.47) | 1.62 (1.41-1.82) |
| **Age range (%\|x, CI 95%)** | | | | | | | | | | | |
| Age<18 | 4.94 (4.88-5.01) | 0.08 (0.06-0.10) | 0.94 (0.88-1.01) | 1.48 (1.38-1.57) | 1.47 (1.24-1.70) | 0.06 (0.00-0.15) | 0.90 (0.84-0.96) | 0.62 (0.51-0.73) | 4.58 (4.25-4.92) | 5.07 (4.26-5.87) | 0.04 (0.01-0.07) |
| Age18-49 | 74.70 (74.56-74.84) | 19.81 (19.53-20.09) | 68.30 (67.99-68.61) | 73.62 (73.27-73.97) | 44.13 (43.17-45.09) | 11.39 (10.27-12.51) | 40.80 (40.50-41.09) | 25.24 (24.61-25.87) | 49.75 (48.95-50.56) | 37.76 (35.97-39.54) | 18.47 (17.85-19.09) |
| Age50-64 | 15.85 (15.73-15.96) | 42.57 (42.22-42.92) | 24.29 (24.00-24.58) | 17.69 (17.39-18.00) | 26.80 (25.94-27.66) | 29.14 (27.54-30.74) | 36.51 (36.22-36.80) | 35.04 (34.35-35.73) | 27.17 (26.45-27.89) | 29.56 (27.88-31.23) | 37.62 (36.84-38.40) |
| Age>64 | 4.51 (4.44-4.57) | 37.55 (37.21-37.89) | 6.47 (6.30-6.63) | 7.21 (7.01-7.41) | 27.60 (26.73-28.46) | 59.41 (57.68-61.14) | 21.79 (21.55-22.04) | 39.10 (38.40-39.81) | 18.49 (17.87-19.12) | 27.62 (25.98-29.27) | 43.87 (43.07-44.66) |
| **Demographics (%\|x, CI 95%)** | | | | | | | | | | | |
| PREGNANT | 1.08 (1.05-1.12) | 0.10 (0.07-0.12) | 0.49 (0.44-0.53) | 0.33 (0.29-0.38) | 0.51 (0.37-0.64) | 0.03 (0.00-0.10) | 0.49 (0.44-0.53) | 0.08 (0.04-0.12) | 1.78 (1.57-1.99) | 0.53 (0.26-0.79) | 0.01 (0.00-0.02) |
| Females | 50.98 (50.82-51.14) | 47.02 (46.67-47.38) | 51.08 (50.75-51.42) | 39.59 (39.21-39.98) | 49.94 (48.97-50.90) | 46.95 (45.19-48.71) | 42.63 (42.33-42.92) | 44.33 (43.61-45.05) | 55.65 (54.85-56.45) | 50.32 (48.48-52.15) | 41.73 (40.94-42.51) |
| Age | 38.10 (38.05-38.14) | 60.32 (60.23-60.41) | 43.36 (43.28-43.45) | 40.65 (40.54-40.77) | 52.73 (52.37-53.09) | 66.73 (66.24-67.21) | 52.72 (52.63-52.82) | 58.90 (58.67-59.14) | 47.74 (47.45-48.03) | 51.74 (51.04-52.45) | 61.90 (61.69-62.12) |
| **Outcomes (%\|x, CI 95%)** | | | | | | | | | | | |
| Recovered | 98.50 (98.46-98.54) | 63.88 (63.54-64.22) | 92.56 (92.39-92.74) | 94.59 (94.41-94.77) | 84.00 (83.29-84.71) | 51.24 (49.48-53.00) | 81.44 (81.20-81.67) | 61.94 (61.24-62.64) | 82.39 (81.77-83.00) | 67.49 (65.77-69.21) | 61.97 (61.20-62.75) |
| Survival days (deceased) | 13.46 (13.21-13.71) | 13.38 (13.27-13.48) | 14.08 (13.85-14.31) | 14.34 (14.01-14.67) | 13.02 (12.57-13.46) | 11.42 (11.02-11.83) | 14.01 (13.88-14.14) | 12.37 (12.16-12.58) | 13.38 (13.01-13.76) | 12.55 (11.91-13.19) | 13.24 (13.01-13.48) |
| Survival>15days | 99.04 (99.01-99.07) | 76.94 (76.64-77.24) | 95.43 (95.29-95.57) | 96.78 (96.64-96.92) | 89.57 (88.98-90.16) | 63.73 (62.04-65.42) | 88.64 (88.45-88.83) | 73.69 (73.06-74.33) | 88.67 (88.16-89.18) | 78.29 (76.77-79.81) | 75.12 (74.43-75.81) |
| Survival>30days | 98.57 (98.54-98.61) | 65.70 (65.36-66.04) | 92.97 (92.80-93.14) | 94.92 (94.75-95.09) | 84.80 (84.11-85.50) | 53.11 (51.36-54.87) | 82.60 (82.37-82.82) | 63.75 (63.05-64.44) | 83.35 (82.75-83.95) | 68.90 (67.19-70.60) | 63.90 (63.13-64.67) |
| Survival>15days_Deceased | 36.34 (35.10-37.58) | 36.17 (35.60-36.74) | 38.61 (37.41-39.81) | 40.54 (38.87-42.22) | 34.77 (32.47-37.08) | 25.61 (23.41-27.81) | 38.78 (38.10-39.46) | 30.88 (29.80-31.96) | 35.68 (33.84-37.52) | 33.23 (30.19-36.26) | 34.57 (33.33-35.80) |
| Survival>30days_Deceased | 5.16 (4.59-5.73) | 5.04 (4.78-5.30) | 5.51 (4.95-6.08) | 6.07 (5.26-6.89) | 4.99 (3.94-6.05) | 3.84 (2.87-4.81) | 6.24 (5.90-6.58) | 4.75 (4.25-5.25) | 5.46 (4.59-6.33) | 4.33 (3.02-5.64) | 5.07 (4.50-5.64) |
| Symptoms to hospitalization days | 4.17 (4.13-4.22) | 5.06 (5.03-5.09) | 5.28 (5.23-5.34) | 5.07 (4.99-5.15) | 4.73 (4.60-4.86) | 4.36 (4.21-4.51) | 4.82 (4.78-4.85) | 4.47 (4.40-4.54) | 4.34 (4.24-4.44) | 4.07 (3.89-4.24) | 5.07 (4.99-5.14) |
| Hospitalized | 6.22 (6.14-6.30) | 67.22 (66.89-67.56) | 19.31 (19.04-19.57) | 14.58 (14.30-14.86) | 32.51 (31.60-33.41) | 73.54 (71.99-75.09) | 44.41 (44.11-44.71) | 64.20 (63.51-64.89) | 38.37 (37.59-39.16) | 61.01 (59.22-62.81) | 67.13 (66.38-67.89) |
| ICU | 0.23 (0.21-0.24) | 7.24 (7.06-7.43) | 1.88 (1.79-1.97) | 1.14 (1.06-1.23) | 2.24 (1.95-2.53) | 6.52 (5.65-7.39) | 4.11 (3.99-4.23) | 4.58 (4.28-4.89) | 2.84 (2.57-3.10) | 5.10 (4.29-5.91) | 6.69 (6.29-7.09) |
| INTUBATED | 0.51 (0.48-0.53) | 14.11 (13.87-14.36) | 3.24 (3.12-3.36) | 2.27 (2.16-2.39) | 5.68 (5.23-6.13) | 15.26 (14.00-16.53) | 7.76 (7.60-7.92) | 12.29 (11.82-12.77) | 6.87 (6.46-7.28) | 12.81 (11.58-14.04) | 14.45 (13.89-15.02) |
| Other case contact | 49.42 (49.26-49.58) | 27.32 (27.00-27.64) | 46.42 (46.08-46.75) | 50.81 (50.41-51.20) | 38.70 (37.76-39.65) | 21.04 (19.60-22.47) | 35.17 (34.88-35.45) | 24.49 (23.87-25.12) | 35.99 (35.21-36.76) | 27.59 (25.94-29.23) | 27.68 (26.96-28.40) |

### 2. Survival days distribution

For completeness, we excluded patients who presented symptoms after September 30 because 95.532% of deceased patients died within 31 days (Figure 1). Some abnormal values are likely to be artificial errors.

**Figure 1**. Distribution of COVID-19 patients' survival days according to the difference between symptoms date and death date.

| Survival weeks | <=4 weeks | <=31 days | <=6 weeks | <= 8 weeks |
|---|---|---|---|---|
| % individuals | 93.111% | 95.284 | 98.7679% | 99.615% |

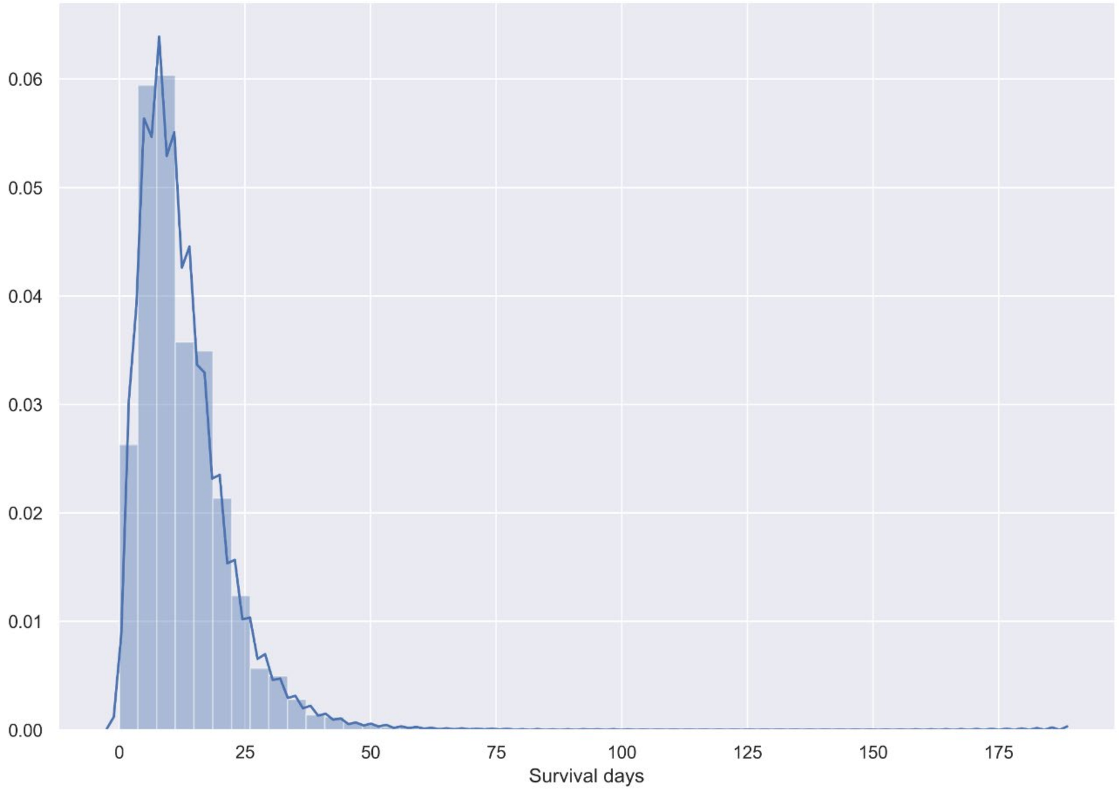

### 3. DQ results regarding temporal and multi-source variability

This section illustrates in more detail about DQ analyses (EHR temporal and source variability[1,2]) taken before the clustering analysis. The dataset's temporal variability assessment displays a variable transient state in some variables' distributions from January to April (Figure 2A), possibly associated to the smaller sample size at these moths (Figure 2B). After April, the variation started to stabilize since hospitalization's probability starts to decrease slightly until October. Afterwards, the number of patients was extremely low in the dataset due to a delayed update of patients' information. By excluding those periods in the temporal analyses, we can observe some changes in the age of patients over time (Figure 2B), as well as in the absolute number of patients regarding age (Figure 2C), and patterns in the entity of medical unit (Figure 2D). Nevertheless, for the meta-clustering analysis all the period was included, given a flat behavior over time in clinical variables.

The temporal variability of all studied variables is available in http://ehrtemporalvariability.upv.es/ and in a specific tutorial notebook at http://personales.upv.es/carsaesi/covid19-metaclustering/EHRTemporalVariability_tutorial.html

**Figure 2.** Temporal heatmap regarding hospitalization and diabetes. Y-axis corresponds with the studied variables, whereas the X-axis corresponds with the date. (A) Probability distribution of the hospitalization. (B) Probability distribution of age. (C) Absolute frequencies of age. (D) Probability distribution of entity of medical unit. Mexico, January 13-November 2, 2020.

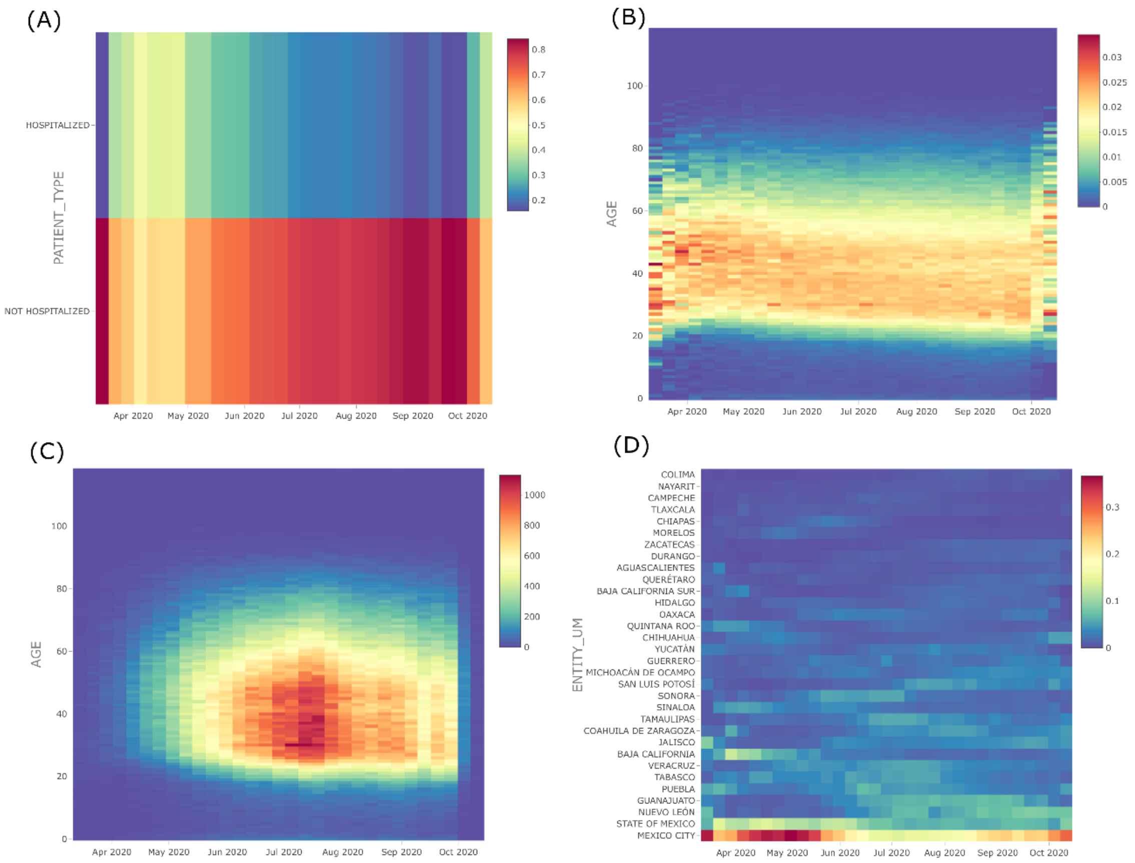

The dataset's multi-source variability assessment found interesting variability patterns in the distributions among data sources in some variables by comparing Mexico's states and the type of clinical institutions where patients received medical attention. Figure 3A shows a striking discrepancy among the states, for example, Hidalgo, Baja California and Morelos all together with Oaxaca, Coahuila de Zaragoza and Durango are located at the left and right extreme, implying these two groups of stats are significantly different upon inclination toward different types of the cluster (in terms of severity, age, and gender) albeit they both are most developed states with similar resources level.

Figure 3B displays the variability between different clinical institution. Surprisingly, we can observe two groups. For example, STATE, SEMAR, UNIVERSITARY, SSA, RED CROSS and SEDENA altogether form one single group and are far located compared with PEMEX, IMSS, ISSSTE, Municipal and the private institution form another group. Surprisingly, most patients were received by the public health system (SSA) and the two main social security systems (IMSS and ISSSTE) whose SPO values differ notably. Thus, after finding these source variabilities, we decided to include all data in the meta-clustering analysis to further evaluate the effect of these sources' differences.

**Figure 3.** MSV plot. Each circle represents a data source, where the distances among them represent the distances among their distributions. The color indicates the SPO of each source, and the circle size the source sample size. (A) State variability. (B) Type of clinical institution variability. The 11 meta-clusters were used as input which means that the larger the distance between two states or type of clinical institution the more different their inclination toward clusters of different severity, age, and gender. Mexico, January 13–September 30, 2020.

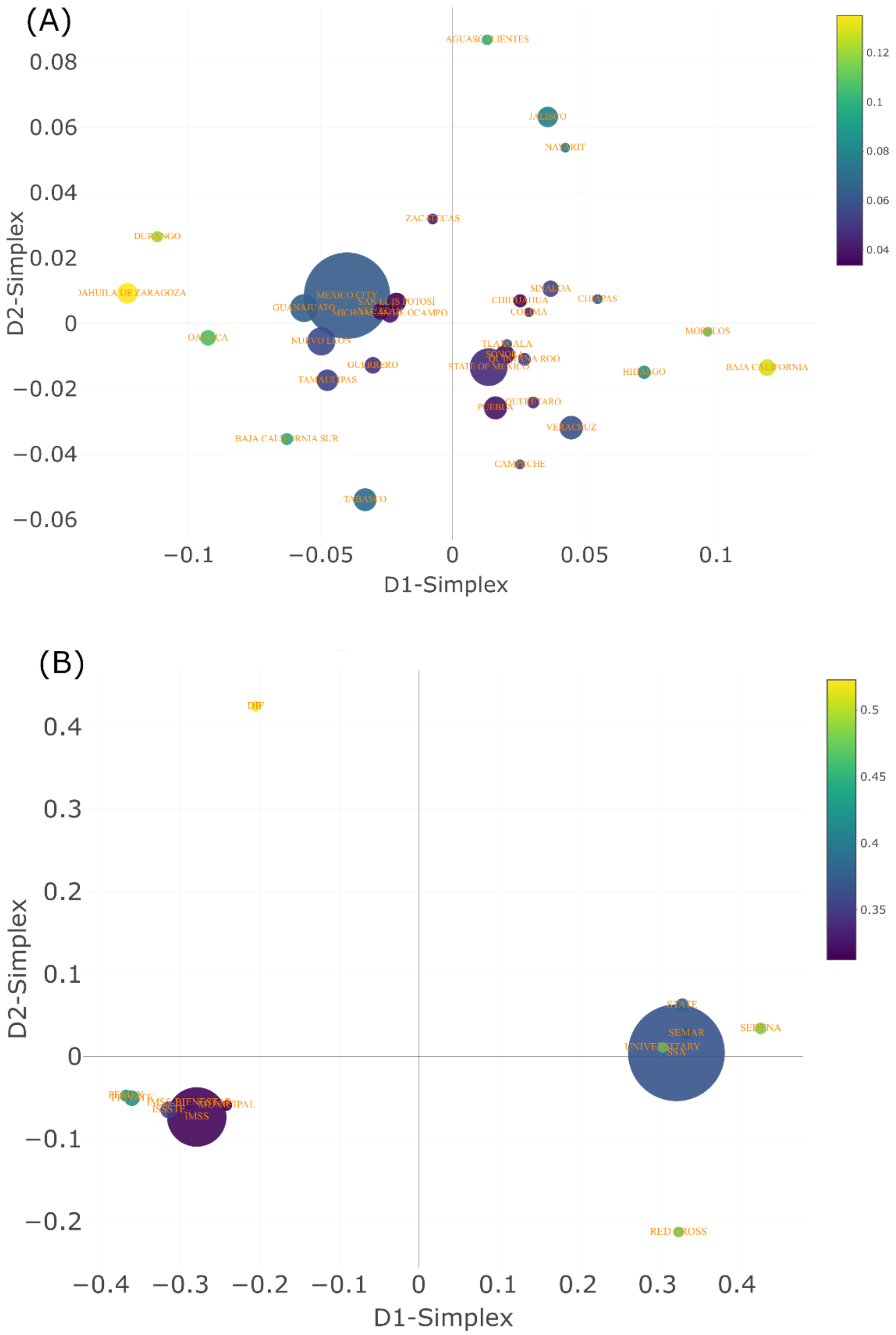

### 4. List of variables contained in the original dataset

Table 2 displays the variables in the original dataset. The dataset is available in our GitHub repository https://github.com/bdslab-upv/covid19-metaclustering

**Table 2.** Variables in the original dataset. English version translated by the authors (original in Spanish). Mexico, January 13-November 2, 2020.

| Variable | Description | Type (value/format) |
|---|---|---|
| Sex | Sex of the person | Discrete (Male, Female) |
| Age | Age in years at the time of the admission | Numerical Integer |
| Pregnant | Presence of pregnancy | Discrete (Yes, No) |
| Obesity | Presence of obesity | Discrete (Yes, No) |
| Smoke | Presence of smoking habit | Discrete (Yes, No) |
| Pneumonia | Presence of pneumonia | Discrete (Yes, No) |
| Diabetes | Presence of diabetes | Discrete (Yes, No) |
| COPD | Presence of chronic obstructive pulmonary disease | Discrete (Yes, No) |
| Asthma | Presence of asthma | Discrete (Yes, No) |
| INMUSUPR | Presence of immunosuppression | Discrete (Yes, No) |
| Hypertension | Presence of hypertension | Discrete (Yes, No) |
| CKD | Presence of chronic kidney disease | Discrete (Yes, No) |
| Cardiovascular | Presence of cardiovascular | Discrete (Yes, No) |
| Other disease | Presence of other diseases | Discrete (Yes, No) |
| Patient type | Whether a patient was hospitalized | Discrete (Hospitalized, Non-hospitalized) |
| Intubated | Whether a patient was intubated | Discrete (Yes, No, Not applied) |
| ICU | Whether a patient had been in an intensive care unit | Discrete (Yes, No, Not applied) |
| Other case contact | Whether a patient was detected to have contacted with other coronavirus cases | Discrete (Yes, No) |
| Sector | Type of institution of the National Health System that provided medical care | Discrete[a] |
| Last_update | The last update of a patient's condition | Date (dd/mm/yyyy) |
| ID_registration | A random ID number of the case | Numerical Integer |
| Entity_um | The state where a patient received attention from medical unit | Discrete |
| Entity_nat | The state where a patient was born | Discrete |
| Entity_res | The resident state of a patient | Discrete |
| Municipality_res | The resident municipality of a patient | Discrete |
| Origin | whether the patient's surveillance was carried out through the system of respiratory disease monitoring units (USMER) | Discrete (USMER, Non-USMER) |
| Country_Origin | The country where a patient came from | Discrete |
| Country_nationality | Patient's nationality | Discrete |
| Nationality | Whether a patient has Mexican nationality | Discrete (Mexican, Foreigner) |
| Migrant | Whether a patient is emigrant | Discrete (Yes, No) |
| Speak_indigenous_language | whether a patient speaks an indigenous language | Discrete (Yes, No) |
| Indigenous | Whether a patient is an indigenous citizen | Discrete (Yes, No) |
| Tested | Whether the patient was tested to coronavirus | Discrete (Yes, No) |
| Result_lab | Coronavirus test result | Discrete (Positive SARS-CoV-2, Non-Positive SARS-CoV-2, Pending, Inadequate result, Not Applied) |
| Final_clasification | Identify which sector confirmed the patient as a coronavirus case | Discrete[b] |
| Admission_date | The date when a patient attended by the care unit (not necessarily hospitalized) | Date (dd/mm/yyyy) |
| Symptoms_data | The date when a patient presented symptoms | Date (dd/mm/yyyy) |
| Death_date | The date when a patient defunct | Date (dd/mm/yyyy) |

[a]*IMSS, SSA, ISSSTE, PRIVATE, PEMEX, STATE, SEMAR, SEDENA, IMSS-BIENESTAR, UNIVERSITARY, MUNICIPAL, RED CROSS, DIF.*

[b]*Confirmed by laboratory test, Negative tested by laboratory, Not tested by laboratory, Invalid by laboratory, confirmed by epidemiol ogical clinical association, confirmed by ruling committee, Suspected case.*

### 5. Dataset descriptive statistics describing age and gender groups difference

This section provides descriptive analyses for the variables and assessed for differences between groups using the following statistics: odds ratio and chi-square (χ2) test for categorical variables, one sample t-test for two normally distributed numerical variables, and One-way ANOVA test when there are more than two means of samples to compare. The normality of two included variables was defined by both visual plot and Shapiro-Wilk test. $P < 0.05$ was considered to be significant.

Table 3 shows the epidemiological characteristics and clinical outcomes of the dataset patients, and the gender difference.

From the 778,692 COVID-19 patients, 402,655 (51.7%) were male, and 376,037 (48.3%) were female with a male-to-female sex ratio of 1:0.93. The patients who aged 18-49 accounted for the largest proportion (60.4%), and the mean age was 44.5±16.7 years. The age-independent mortality rate was 10.5% (93.3% and 90% survived more than 15 and 30 days respectively); whereas among deceased patients, 36.34% and 5.38% survived more than 15 and 30 days respectively. Furthermore, the mean days for a hospitalized patient from presenting the symptoms to hospitalization was 4.8±3.7 days (23.5% were hospitalized).

There is a baseline significant severity difference between male and female. For example, 21.3% of male presented pneumonia with a mortality of 13.1%, whereas women presented 14.6% pneumonia rate and 7.9% mortality rate (Odds Ratio: 1.58 [95%CI; 1.56-1.60] and 1.76 [95%CI; 1.74-1.79] respectively, male vs female). COPD and hypertension showed no significant difference. Noteworthy, large sample sizes contribute to test positive for statistically significant differences while reducing confidence intervals.

**Table 3.** Epidemiological characteristics and demographic features of 778,692 COVID-19 patients. *P* was calculated for normally distributed numerical variables using one sample T-test and odds ratio for categorical variables. Mexico, January 13–September 30, 2020.

| | Total (n = 778,692) | Male (n = 402,655) | Female (n = 37,6037) | *P* /Odds Ratio (95%CI) (Male vs Female) |
|---|---|---|---|---|
| Age, mean (SD) | 44.5 (16.7) | 45.2 (16.8) | 43.8 (16.5) | *P*<0.001[a] |
| Age range, n (%) | | | | |
| <18 | 22,868 (2.9) | 11,572 (2.9) | 11,296 (3.0) | OR=0.96 [0.93-0.98] |
| 18-49 | 470,349 (60.4) | 236,765 (58.8) | 233,584 (62.1) | OR=0.87 [0.86-0.88] |
| 50-64 | 184,452 (23.7) | 97,943 (24.3) | 86,509 (23.0) | OR=1.07 [1.06-1.08] |
| >64 | 101,023 (13.0) | 56,375 (14.0) | 44,648 (11.9) | OR=1.21 [1.19-1.22] |
| Pregnant, (%) | 5,735 (0.7) | 0 (0) | 5735 (1.5) | |
| Habits, (%) | | | | |
| Smoke | 56,960 (7.3) | 39,355 (9.8) | 17,605 (4.7) | OR=2.21 [2.17-2.25] |
| Obesity | 138,929 (17.8) | 68,858 (17.1) | 70,071 (18.6) | OR=0.90 [0.89-0.91] |
| Comorbidities, n (%) | | | | |
| Diabetes | 118,867 (15.3) | 62,495 (15.5) | 56,372 (15.0) | OR=1.04 [1.03-1.05] |
| COPD | 11,119 (1.4) | 5,677 (1.4) | 5,442 (1.4) | OR=0.97 [0.94-1.01] |
| Asthma | 20,057 (2.6) | 7,708 (1.9) | 1,2349 (3.3) | OR=0.57 [0.56-0.59] |
| INMUSUPR | 8,311 (1.1) | 3,939 (1.0) | 4,372 (1.2) | OR=0.84 [0.80-0.88] |
| Hypertension | 149,444 (19.2) | 76,966 (19.1) | 72,478 (19.3) | OR=0.99 [0.98-1.00] |
| CKD | 14,526 (1.9) | 8,234 (2.0) | 6,292 (1.7) | OR=1.23 [1.19-1.27] |
| Cardiovascular | 15,072 (1.9) | 8,526 (2.1) | 6,546 (1.7) | OR=1.22 [1.18-1.26] |
| Other disease | 18,525 (2.4) | 8,232 (2.0) | 10,293 (2.7) | OR=0.74 [0.72-0.76] |
| Treatment, n (%) | | | | |
| Hospitalized | 182,675 (23.5) | 110,758 (27.5) | 71,917 (19.1) | OR=1.60 [1.59-1.62] |
| Intubated | 31,978 (4.1) | 20,680 (5.1) | 11,298 (3.0) | OR=1.75 [1.71-1.79] |
| ICU | 15,916 (2.0) | 10,156 (2.5) | 5,760 (1.5) | OR=1.66 [1.61-1.72] |
| Pneumonia | 140,720 (18.1) | 85,692 (21.3) | 55,028 (14.6) | OR=1.58 [1.56-1.60] |
| Mortality, n (%) | 82,077 (10.5) | 52,553 (13.1) | 29,524 (7.9) | OR=1.76 [1.74-1.79] |
| Survival>15days, n (%) | 726,443 (93.3) | 369,553 (91.8) | 356,890 (94.9) | OR=0.60 [0.59-0.61] |
| Survival>30days, n (%) | 701,027 (90.0) | 352,913 (87.6) | 348,114 (92.6) | OR=0.57 [0.56- 0.58] |
| Survival>15days_deceased, n (%) | 29,828 (36.34) | 19,451 (37.01) | 10,377 (35.15) | OR=1.08 [1.05-1.12] |
| Survival>30days_deceased, n (%) | 4,412 (5.38) | 2,811 (5.35) | 1,601 (5.42) | OR=0.99 [0.93-1.05] |
| From Symptom to Hospital days, mean (SD) | 4.8 (3.7) | 4.9 (3.7) | 4.7 (3.6) | *P*<0.001[a] |
| Other case contact, n (%) | 338,709 (43.5) | 161,772 (40.2) | 176,937 (47.1) | OR=0.76 [0.75-0.76] |

[a]*One-sample T-test.*

Table 4 shows the discrepancy in studied characteristics among different age-groups (<18, 18-49, 50-64, and >64). It shows asthma and pregnancy are prone to young people, whereas the smoke rates are similar between adults of different ages. Older patients have clearly higher morbimortality, comorbidity and clinical care rate since age and gender are highly correlated with comorbidity and habit (e.g., smoke and obesity).

**Table 4.** Age differences in regard to epidemiological characteristics and features by COVID-19 subgroups. Mexico, January 13–September 30, 2020.

| | ≤17 (n=22,868) | 18-49 (n=470,349) | 50-64 (n=184,452) | ≥65 (n=101,023) | *P* |
|---|---|---|---|---|---|
| Age, mean (SD) | 10.5 (5.5) | 35.4 (8.3) | 56.2 (4.2) | 73.4 (7.0) | <0.001[a] |
| Sex, n (%) | | | | | <0.001[b] |
| Female | 11,296 (49.4) | 233,584 (49.7) | 86,509 (46.9) | 44,648 (44.2) | |
| Male | 11,572 (50.6) | 236,765 (50.3) | 97,943 (53.1) | 56,375 (55.8) | |
| Features (%) | | | | | |
| Pregnant | 133 (0.6) | 5,587 (1.2) | 10 (0.0) | 5 (0.0) | <0.001[b] |
| Smoke | 187 (0.8) | 37,256 (7.9) | 11,880 (6.4) | 7,637 (7.6) | <0.001[b] |
| Obesity | 974 (4.3) | 80,585 (17.1) | 39,797 (21.6) | 17,573 (17.4) | <0.001[b] |
| Comorbidities, n (%) | | | | | |
| Pneumonia | 1,223 (5.3) | 44,070 (9.4) | 49,514 (26.8) | 45,913 (45.4) | <0.001[b] |
| Diabetes | 158 (0.7) | 31,405 (6.7) | 50,360 (27.3) | 36,944 (36.6) | <0.001[b] |
| COPD | 19 (0.1) | 1,557 (0.3) | 3,208 (1.7) | 6,335 (6.3) | <0.001[b] |
| Asthma | 771 (3.4) | 13,009 (2.8) | 4,267 (2.3) | 2,010 (2.0) | <0.001[b] |
| INMUSUPR | 401 (1.8) | 3,335 (0.7) | 2,617 (1.4) | 1,958 (1.9) | <0.001[b] |
| Hypertension | 132 (0.6) | 3,9577 (8.4) | 58,807 (31.9) | 50,928 (50.4) | <0.001[b] |
| CKD | 107 (0.5) | 4,693 (1.0) | 5,031 (2.7) | 4,695 (4.6) | <0.001[b] |
| Cardiovascular | 197 (0.9) | 3,549 (0.8) | 4,573 (2.5) | 6,753 (6.7) | <0.001[b] |
| Other disease | 578 (2.5) | 8,548 (1.8) | 5,091 (2.8) | 4,308 (4.3) | <0.001[b] |
| Treatment, n (%) | | | | | |
| Hospitalized | 2,572 (11.2) | 55,022 (11.7) | 64,583 (35.0) | 60,498 (59.9) | <0.001[b] |
| Intubated | 344 (1.5) | 7,029 (1.5) | 12,167 (6.6) | 12,438 (12.3) | <0.001[b] |
| ICU | 485 (2.1) | 4,521 (1.0) | 5,727 (3.1) | 5,183 (5.1) | <0.001[b] |
| Mortality, n (%) | 276 (1.2) | 14,042 (3.0) | 29,485 (16.0) | 38,274 (37.9) | <0.001[b] |
| Survival days, mean (SD) | 11.9 (11.1) | 13.4 (9.4) | 14.0 (9.5) | 13.1 (8.9) | <0.001[a] |
| Survival<15days, n (%) | 204 (0.9) | 8,941 (1.9) | 18,151 (9.8) | 24,953 (24.7) | <0.001[b] |
| Survival<30days, n (%) | 256 (1.1) | 13,303 (2.8) | 27,692 (15.0) | 36,414 (36.0) | <0.001[b] |
| From Symptom to Hospital days, mean (SD) | 2.2 (2.9) | 4.7 (3.6) | 5.0 (3.7) | 4.8 (3.7) | <0.001[a] |
| Other case contact, n (%) | 13,411 (58.6) | 228,647 (48.6) | 69,495 (37.7) | 27,156 (26.9) | <0.001[b] |

[a]*One-way ANOVA.*

[b]*Pearson Chi-square Goodness of Fit test.*

### 6. A brief analysis about the pregnancy association in female patients

A total of 5,735 pregnant patients (0.74% of total cases) were included in our studied sample. Pregnant women were primarily aged between 15-45 (n = 5,697, 99.34% of total); so we decided to quantify the mortality and severity difference between pregnant and non-pregnant women (n = 410,643) in this age range. Figure 4 demonstrates that pregnancy is associated with mortality significantly (p<0.001) with an increase in the incidence of pneumonia, INMUSUPR, and other diseases rate (p<0.001). Since pregnant women experience immunologic and physiologic alteration that may increase their risk for more severe illness from respiratory infections[3,4,5]. Conversely, the incidence of CKD, asthma, smoke and obesity were lower (p≤0.003). Interestingly, pregnancy are not associated with the prevalence of cardiovascular and only a slight increase in diabetes was found; this is contrary to a recent report in USA[3] that mentioned a double rate of diabetes and cardiovascular in pregnant women.

**Figure 4.** Probability between pregnant and non-pregnant COVID-19 patients (who aged between 15 to 45; grouped bins in groups of 5 years) in obesity, smoking habit, comorbidities and other case contact. Chi-Square test was used. Mexico, January 13–September 30, 2020.

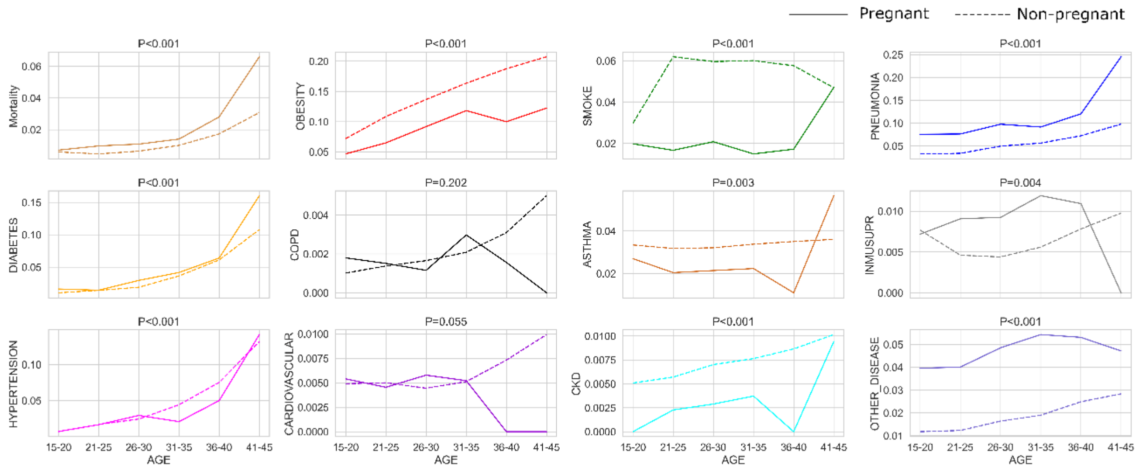

#### 7. Number of individual among age-gender specific clusters

Table 5. Number of cases at each of the 56 age-gender specific clusters. Mexico, January 13–September 30, 2020.

| Cluster size (n) | 0-1,000 | 1,001-5,000 | 5,001-10,000 | 10,001-30,000 | >=30,000 |
|---|---|---|---|---|---|

| Cluster ID (unrelated among groups) | <18M | <18F | 18-49M | 18-49F | 50-64M | 50-64F | >64M | >64F |
|---|---|---|---|---|---|---|---|---|
| 1 | 9,463 | 753 | 138,266 | 149,573 | 17,716 | 29,725 | 16,997 | 14,861 |
| 2 | 542 | 10,228 | 19,863 | 22,897 | 3,525 | 4,017 | 8,057 | 2,806 |
| 3 | 410 | 81 | 24,449 | 17,555 | 41,583 | 15,293 | 3,372 | 11,023 |
| 4 | 969 | 234 | 3,947 | 6,776 | 12,113 | 3,478 | 17,144 | 6,342 |
| 5 | 188 | | 22,905 | 21,131 | 10,231 | 4,042 | 1,192 | 1,886 |
| 6 | | | 22,224 | 12,186 | 2,543 | 16,527 | 4,334 | 2,469 |
| 7 | | | 5,111 | 3,466 | 484 | 2,570 | 2,866 | 4,001 |
| 8 | | | | | 2,884 | 10,857 | 2,413 | 1,260 |
| 9 | | | | | 6,864 | | | |
